# A unified modeling platform for informing cervical cancer prevention policy decisions in 132 low- and middle-income countries

**DOI:** 10.64898/2026.03.18.26348700

**Authors:** Irene Man, Alina Macacu, Mattis Eynard, Indira Adhikari, Andrea Gini, Damien Georges, Iacopo Baussano

## Abstract

**Background:** Public health decision modelling tools designed to inform cervical cancer prevention policies in low- and middle-income countries (LMICs) are useful but scarce. Important challenges herein are the often missing or inconsistently collected cervical cancer epidemiological data, and the lack of a systematic approach to deal with such data limitations.

**Methodology/Principal Findings:** We developed a unified modelling platform and workflow to enable cervical cancer modelling in 132 LMICs based on the previously developed “footprinting” approach, through the following steps: 1) With sexual behavior data from the Demographic Health Surveys (DHS), which were available for a large number of LMICs (70/132), we identified clusters of countries which represent distinct patterns of human papillomavirus (HPV) transmission. The 7 resulting clusters correspond to a gradient of HPV prevalence and cervical cancer risk and exhibit clear geographical separation. 2) The remaining LMICs were classified into the identified clusters based on geographical proximity so that each LMIC was grouped to a cluster. Goodness of classification was validated using available epidemiological data. 3) We then calibrated the HPV transmission and cervical cancer progression models of the IARC/WHO METHIS platform to the 132 LMICS, first by cluster then by country, using the available data on sexual behavior (from DHS), HPV prevalence (from literature search), and cervical cancer incidence (from GLOBOCAN).

**Conclusions/Significance:** A unified workflow and platform designed by IARC/WHO for public health decision modelling of cervical cancer prevention in 132 LMICs is now available. It is ready to be used to support global and local stakeholders to coordinate, design, and implement impactful and efficient prevention policies and will help to accelerate cervical cancer elimination.

## Introduction

Cervical cancer, despite being preventable, is still an important public health problem, especially in low-resource settings (1). In 2022, the incidence of cervical cancer in most low- and middle-income countries (LMICs) was still far above the elimination target (of 4 cases per 100,000 women per year) set by the World Health Organization (WHO): 25.4, 16.9, and 14.2 cases in low-, lower-middle-, and upper-middle-income countries, respectively (1).

A powerful yet under-utilized tool to support global and local public health stakeholders in cervical cancer prevention and control is public health decision modeling (2, 3). Modeling enables the quantification of disease burden that is expected and preventable through public health interventions as well as the broader societal and economic benefits. This information is key for justifying past investment in prevention programs and their continuation in the future. Modeling also allows comparison of different alternative policies, which help to design and implement impactful and efficient prevention programs adapted to the local needs and available resources.

Over time, various research groups, including ours, have developed public health decision models for cervical cancer to many countries (4, 5); however, only a small number of these models have been specifically designed for LMICs and provide results for multiple LMICS in a comparative way (6-8). An important challenge herein is the paucity of high-quality and consistently collected epidemiological data to parametrize the models (e.g., data on sexual behavior, HPV prevalence, and cervical cancer), and the lack of a robust theoretical framework to utilize such limited data. To overcome these challenges, we have previously developed the so-called “Footprinting” framework, which provides a workflow for modeling a large number of countries/regions while accounting for the heterogeneous and fragmented data (9). This framework has already been successfully applied to model large countries with many subnational regions, such as India and Brazil, and has generated useful modelling results informing local cervical cancer elimination polices (10-14).

In this paper, following the same Footprinting framework (9), we constructed a unified modelling workflow to inform cervical cancer prevention policies in 132 LMICs. To do so, we first identified clusters of LMICs representing different archetypes of epidemiological patterns and subsequently calibrated the IARC/WHO METHIS cervical cancer modelling platform (15) to each cluster and country.

## Results

### A unified public health decision modelling workflow for cervical cancer prevention in LMICs

Following the Footprinting framework we proposed previously (9), we constructed a unified workflow for modelling cervical cancer prevention in 132 LMICs, through the following steps: *clustering, classification, and calibration*.

#### Clustering step

We identified the main archetypes/patterns of sexual behavior and the corresponding clusters of LMICs. Sexual behavior data from Demographic and Health Surveys (DHS) were used for this purpose (16), as DHS collected data on many sexual behavior indicators in a standardized way in many countries. From the 132 LMICs, there were 70 countries with sufficient DHS data available for the clustering analysis. Note that we found HPV prevalence data less adequate for the clustering step, as these are generally less consistently measured and collected. We also did not use cervical cancer incidence. We found it less adequate for identifying clusters which will be used to calibrate the HPV transmission model, as cervical cancer incidence is also modified by the background screening rates.

#### Classification step

We classified the remaining 62 LMICs to the identified clusters. Classification was based on geographical proximity and subsequently validated using the partially available DHS sexual behavior data or GLOBOCAN cervical cancer incidence data (1).

#### Calibration step

We calibrated the models of the IARC/WHO METHIS modelling platform (15). We first pre-calibrated the platform’s HPV transmission model (RHEA), at the cluster level, using cluster-aggregated data of sexual behavior from DHS (16) and HPV prevalence from literature search (see Section 1 of Supplementary material A for details of the search). Subsequently, country-level finetuning was done using country-specific cervical cancer incidence and mortality data from GLOBOCAN (1) in the platform’s cervical cancer progression model (ATLAS). Once calibrated, the models can be used to project expected impact of a range of primary and secondary cervical cancer prevention policies.

See eTable 1 for the list of the 132 included LMICs and the corresponding data availability in terms of sexual behavior, HPV prevalence, and cervical cancer incidence.

In the following sections, we report the detailed results of each step.

### Step 1 – Clustering LMICs based on sexual behavior data

#### Pre-processing using Principal Component Analysis

To ensure the robustness of the clustering analysis, we first pre-processed the input sexual behavior data through a Principal Component Analysis (PCA). This reduced the original 48 indicators into six main dimensions, which together explained 70% of the total variance. The six dimensions to be included in subsequent clustering analysis was determined based on the elbow method (eFigure 3). Each dimension corresponds to a different set of original indicators:

1. Age of first sexual intercourse (women), age of first marriage (women), and age of first childbirth (women)
2. Age of first sexual intercourse (men), lifetime number of sexual partners (men and women), and higher-risk sex (men and women)
3. Age difference between sexual partners, and age of first marriage (men)
4. HIV prevalence and coverage of ART (men and women)
5. Paid sexual intercourse (men), and concurrent sexual partners (men)
6. Number of wives (men), lifetime number of sexual partners (women), and concurrent sexual partners (women)

See eFigures 1 and 2 for the detailed PCA results.

#### Sexual behavior clusters of LMICs identified

Clustering was done using a Gaussian Mixture Model (GMM). We explored results with 4 or 5 clusters, as we considered 3 clusters too few for the number of countries included, and the model no longer converged with more than 5 clusters.

The results with 4 or 5 clusters were both satisfactory, with sufficient visual separation in the included dimensions, especially the first two dimensions (eFigure 4). The final country groupings were similar (eFigure 5). Moving from 4 to 5 clusters mainly led to the split of one cluster, while the other clusters remained mostly intact (eFigures 5 and 6). The two clusters resulting from the split correspond roughly to the countries in America and central Africa. Overall, we found the 4-clustering more adequate for our purpose, as it exhibits clearer geographical separation, with only a few exceptions (Burundi, Rwanda, Comoros, and Nepal). The goodness of the clustering was further demonstrated by the high membership probabilities of >95% for 63 of the 70 LMICs considered (see eTable 3 for the membership probabilities results).

The four main clusters A-D (ordered from top to bottom in eFigure 6) have the following characteristics (see also eTable 4 for the cluster mean of each indicator):

- Cluster A, besides having the ***highest number of lifetime partners***, also stands out due to the ***high HIV prevalence***. This cluster is exclusively located in southern Africa. This cluster is also characterized by ***early first sexual intercourse*** but ***late first marriage***.
- Cluster B does not have high HIV prevalence, but still a ***high number of lifetime partners*** as well as ***early first sexual intercourse***. This cluster has countries around central Africa and America.
- Cluster C is characterized by a ***low number of sexual partners, small age difference between partners***, and the ***latest first sexual intercourse in women***. This cluster has countries in southeastern and central Asia, and Europe.
- Cluster D also has a ***low number of lifetime sex partners***, but ***large age difference between partners*** and ***early marriage in women***, with countries in northern Africa and southern Asia.

To obtain a final clustering with enough granularity while still respecting the geographical distribution, we constructed the final clustering by splitting the 4 main clusters into 7 region-specific subclusters based on separation of continents (Figure 1, eFigure 6). Burundi, Rwanda, Comoros, and Nepal were reassigned to the closest region-specific subclusters. See eTable 5 for the subcluster means of each sexual behavior indicator. The subclusters are ordered by decreasing mean cervical cancer incidence (eFigure 7).

**Figure 1.**
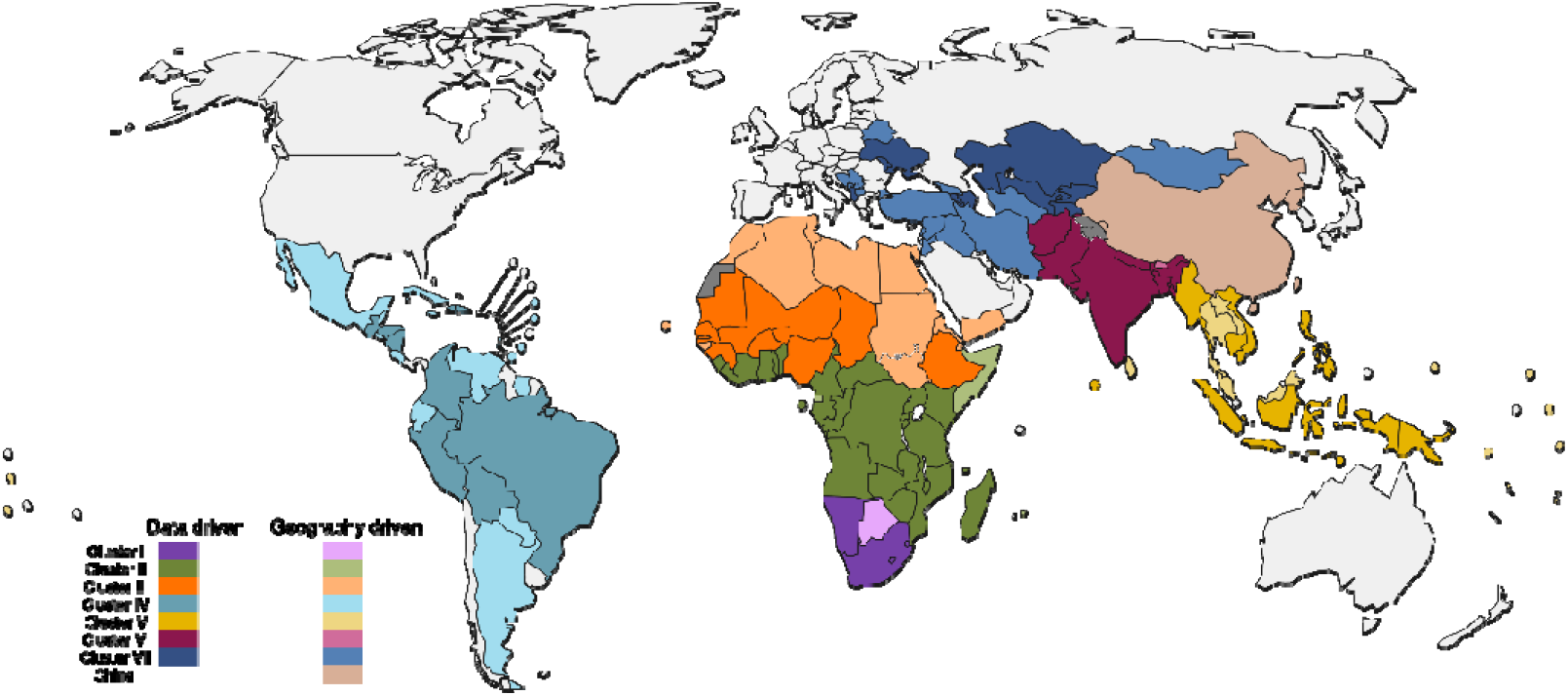
Geographical distribution of the seven region-specific subclusters. The seven colors (purple, green, orange, light blue, yellow, red, and blue) indicate the seven clusters. The darker shade corresponds to countries included in the clustering analysis driven by DHS sexual behavior data. The lighter shade corresponds to countries classified into the identified clusters based on geography.

### Step 2 – Classification of remaining LMICs to identified clusters based on geography

We then classified 60 of the 62 remaining LMICs into the 7 region-specific subclusters, based on proximity to maintain the convenient geographical distribution. China and North Korea were not classified to any cluster. See countries filled with light shades in Figure 1 and eTable 6.

Goodness of classification was validated with the partially available (<50% indicators) DHS sexual behavior data. When unavailable, GLOBOCAN cervical cancer incidence data were used. For nine countries (mostly island nations), no data was available. For the 15 countries with partially available sexual behavior data, we estimated the membership probabilities using the fitted GMM for validation. For 14 countries (i.e., 93%), the geography-driven classified cluster had the highest or second highest probability, demonstrating goodness of fit (eTable 7). Among the remaining 36 countries with GLOBOCAN cervical cancer incidence data, for 21 countries (i.e., 58%), the geography-driven classified cluster has median incidence closest or second closest to the country’s incidence, and for 28 countries (78%), among the first three closest, demonstrating satisfactory goodness of fit (eTable 8).

### Step 3 – Calibration of HPV transmission and cervical cancer progression models

We then calibrated the HPV transmission model (RHEA) of the METHIS platform to sexual behavior data extracted from the DHS (eFigures 8 and 9) and to HPV prevalence data from literature (Figure 2). The calibration was done separately for each cluster, using cluster-aggregated weighted averages. The amount of data available for calibration varied across clusters (Table 1). The percentage of countries with available sexual behavior data to contribute for each cluster ranges from 20% to 67%. For HPV prevalence data, the percentage ranges from 11% to 83%. Some countries had age- and type-specific HPV prevalence data from more than one study, and most studies reported HPV 16 prevalence, with fewer studies reporting prevalence for other high-risk HPV.

**Figure 2.**
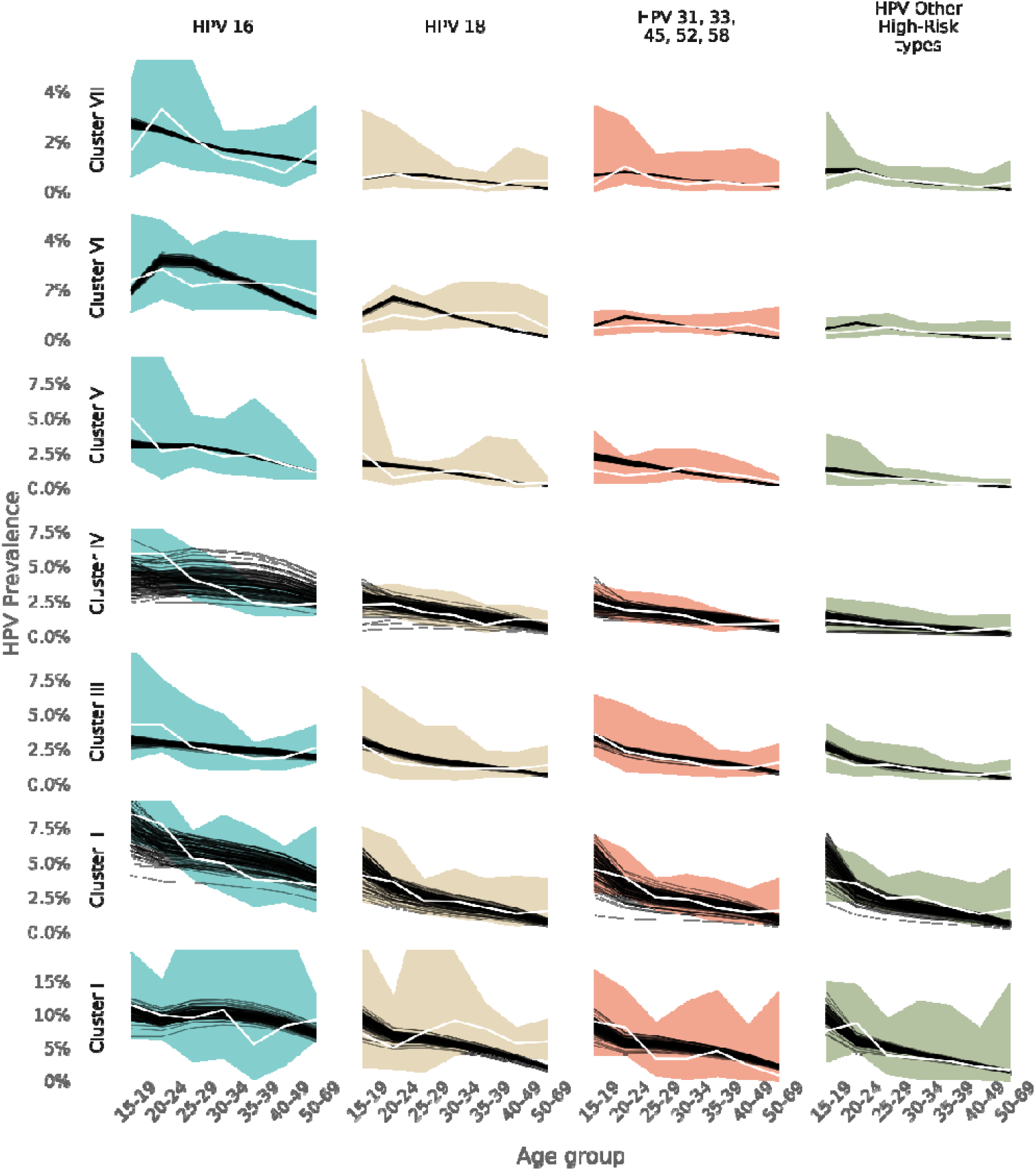
Cluster-level calibration of the HPV transmission model. Best 100 parameters of RHEA calibrated to each cluster, to match the observed cluster aggregated age-specific HPV prevalence (white lines represents the average, while the coloured intervals represents the 95% uncertainty intervals). The calibration was done separately for HPV16, HPV18, HPV 31, 33, 45, 52, 58, and HPV -other HR types. These four groupings of HPV genotypes were modelled separately inside RHEA.

**Table 1.** Data points available for calibration of RHEA the HPV transmission model. **\*** Not all sexual behaviour data needed for calibration was available for each country with DHS data in cluster I. For each study with HPV prevalence data available, up to seven data-points were included, one for each age-group (15-19, 20-24, 25-29, 30-34, 35-39, 40-49, 50-69).

| Cluster | N° of countries in cluster | Sexual behaviour calibration data availability | HPV prevalence calibration data availability |  |  |  |
| --- | --- | --- | --- | --- | --- | --- |
|  |  |  | HPV 16 | HPV 18 | HPV 31, 33, 45, 52, 58 | HPV Other high risk |
| I | 5 | 1 to 3 countries * | 3 countries (5 studies) | 2 countries (4 studies) | 1 country (2 studies) | 1 country (2 studies) |
| II | 27 | 5 countries | 5 countries (8 studies) | 5 countries (8 studies) | 5 countries (8 studies) | 5 countries (8 studies) |
| III | 22 | 7 countries | 7 countries (9 studies) | 7 countries (9 studies) | 7 countries (9 studies) | 7 countries (9 studies) |
| IV | 25 | 7 countries | 7 countries (18 studies) | 7 countries (17 studies) | 7 countries (18 studies) | 7 countries (16 studies) |
| V | 21 | 5 countries | 5 countries (8 studies) | 5 countries (8 studies) | 5 countries (9 studies) | 5 countries (9 studies) |
| VI | 6 | 5 countries | 5 countries (18 studies) | 5 countries (16 studies) | 5 countries (14 studies) | 5 countries (13 studies) |
| VII | 26 | 5 countries | 5 countries (9 studies) | 5 countries (8 studies) | 5 countries (8 studies) | 5 countries (8 studies) |
| Total | 132 | 37 countries | 37 countries (75 studies) | 36 countries (70 studies) | 35 countries (68 studies) | 35 countries (65 studies) |

As the clusters were ordered by cervical cancer incidence, the observed aggregated age- and type-specific HPV prevalence was highest for Cluster I, and lowest for Cluster VII, as expected. In addition, there was a decreasing trend with increasing age (Figure 2). The 100 best fitting parameters identified through calibration (eFigure 10) generally produced HPV prevalence that fitted well the observed age-specific curve within its 95% uncertainty intervals.

## Discussion

In this paper, we constructed a unified modeling workflow and platform to support public health decisions on cervical cancer prevention policies for 132 LMICs, based on the models of the IARC/WHO METIHS platform (15) and calibrated with local data. Using the constructed platform, it is then possible to evaluate and project the impact of prevention strategies to progress towards cervical cancer elimination.

This study has many strengths. We believe that our work has considerably improved the existing set of tools for multi-country modelling in LMICs. It made use of a large amount of data from comprehensive reference databases of sexual behavior and cervical cancer risk (16, 18), and HPV prevalence data from literature that complements previous elaborate systematic reviews (19, 20). Upon gathering a large amount of data, we designed a workflow that makes use of as much data from as many countries as possible. Furthermore, by reporting each step of the workflow in detail, it is reproducible and adaptable for further improvement.

Our two-step approach of cluster-level pre-calibration then country-specific finetuning has various advantages. First, the cluster-level calibrated model is sufficient for answering many decision questions while requiring lower computational effort. It could be used to set up rules of thumb for decisions, e.g., maximum or range of acceptable price for new vaccines for pooled procurement, and prioritization of vaccine target population. Such analyses are helpful for global stakeholders for coordination of global public health actions and for local stakeholders to take certain high-level decisions that required less precise planning. The cluster-level pre-calibration then provides a solid starting point for rapid country-specific finetuning. For instance, more in-depth analyses could be required to inform local stakeholders on detailed planning, for example, exact estimates on resource needed for a prevention program (e.g., budget, vaccines, and healthcare workers).

Another merit of this study is that it has generated a deeper understanding of the patterns of sexual behavior among LMICs. Compared to previous analyses that were more descriptive and mostly emphasize the heterogeneity and complexity of sexual behavior patterns (21), our results provide an pragmatic summary and mapping of the archetypical patterns that is directly usable in a workflow for modeling the HPV transmission in all LMICs. In fact, the same results and methodology are also applicable for other sexually transmitted infections.

Various methodological limitations of this study required discussion. First, while, to our knowledge, we are making the best use of the existing data, there are various limitations with data availability, consistency, and quality. Notably, there are regions (e.g., northern Africa and middle east, island nations, or in conflict-prone and hard-to-reach areas) with extremely limited amounts of data. There are many countries with no DHS sexual behavior data, and among these countries, their cervical cancer incidence and mortality data are often based on extrapolation and estimation instead of primary data. Hence, modeling results for such countries (both by us and other modelling groups) should be interpreted with great care. To overcome this limitation, we need to sustain and expand existing data collection initiatives such as DHS for sexual behavior and other demographic information, IARC GLOBOCAN, which coordinate and train cancer registries for collection of cancer data (18), as well as initiatives to expand and standardize the collection of HPV prevalence data such as Labnet (22) and CHRONOS (23). As another limitation, in the clustering step, after identifying main clusters based on the sexual behavior data, we used geographical distribution to create more refined smaller subclusters. This approach, while not completely data driven, was necessary to safeguard interpretability.

In conclusion, a unified workflow and platform for public health modelling of cervical cancer prevention in 132 LMICs is now ready to be used. It has the potential to accelerate cervical cancer elimination by supporting global and local stakeholders in coordinating, designing, and implementing impactful and efficient prevention policies.

## Materials and Methods

### IARC/WHO METHIS modelling platform

Over time, the Public Health Decision Science Team of IARC/WHO has developed the cervical cancer prevention modelling platform, called METHIS. It consists of two HPV transmission and three cervical cancer progression models, which can be used stand-alone or in combination. Which model or models to use is a trade-off between the availability/quality of informative data and the complexity of the public health questions of interest. The modeling platform can be used to estimate and predict the burden of cervical cancer and the impact of cervical cancer preventive interventions.

Here, we present the unified modeling workflow adapting the METHIS modelling platform’s HPV transmission model (RHEA) and cervical cancer progression model (ATLAS) to 132 LMICs. See the platform website for a detailed description and previous publications using each platform model (15).

### Step 1. Clustering LMICs based on sexual behavior data

The first step of the unified modelling workflow consisted of clustering 132 LMICs according to the main underlying patterns of sexual behaviour that drive the HPV prevalence in each country and ultimately impact the cervical cancer burden in the country.

#### DHS sexual behavior indicators used for clustering

To identify clusters of LMICs, we used data from DHS on 45 indicators related to sexual behavior, which take form as country aggregated mean or proportion. For each country and indicator, we used data from the most recent survey. The included indicators were: age difference between sexual partners (5 indicators), age of first sexual intercourse (12), age of first marriage (12), age of first childbirth (5), lifetime number of sexual partners (2), concurrent sexual partners (2), paid sexual intercourse (2), higher-risk sex (2), condom use during higher-risk sex (2), and number of wives (1). As human immunodeficiency virus (HIV) infection and acquired immunodeficiency syndrome (AIDS) are important risk factors of HPV infection (24), we also included three indicators related to HIV/AIDS: HIV prevalence in the population aged 15+ and the coverage of ART. We used the 2021 UNAIDS estimates (25). For simplicity, we refer to all included data as sexual behavior data in the remainder of the paper. Details of the indicators can be found in eTable 2 and see Supplementary material B for the extracted data.

#### Multiple imputation of missing DHS sexual behavior data

There were 87 countries with at least some DHS data on the selected 45 indicators. Overall, approximately 20% of the data was missing from the dataset. The percentage of missing data ranges from 0% to 76% across countries. To avoid biased results, we included in the clustering analysis only the 72 countries with more than 50% of the indicators available. Of these, 70 are currently LMICs and two (Guyana and Trinidad and Tobago) were LMICs in the past (26). Missing data was imputed through multiple imputations. We assume that data were missing at random and imputed the missing data 500 times by multiple imputation with a predictive mean matching algorithm. The MICE package of R was used (version 3.15.0) (27).

#### Clustering analysis

The high number of indicators and the high correlation between them generally complicates the clustering analysis. Hence, we first conducted a Principal Component Analysis (PCA). The package FactoMineR of R was used (version 2.8) (28).

The PCA-transformed data was then used for clustering with a GMM, which allows for clusters of different shapes and different densities. The package MGMM of R was used (version 1.0.0) (29). Likelihood of a given country belonging to a given cluster was estimated based on probabilities of membership. During the step of multiple imputation, multiple datasets were generated, resulting in multiple representations of each country. To determine the final cluster assignment for a country, we computed the average probability of membership across multiple representations.

### *Step 2. Classification of* remaining LMICs to identified clusters based on geography

#### Validation of geography-driven classification using partially available DHS data

For the 15 countries with some DHS sexual behavior data available (but for <50% indicators), we used the membership probabilities estimable from the fitted GMM for validation. In this, we only used the following indicators, as these were available for most countries: age of first marriage in women (available for all included countries), age of first childbirth (only missing for El Salvador), HIV prevalence in 15-19 years old and coverage of ART (only missing for Jordan, Turkey, and Turkmenistan). Again, missing values were imputed and PCA performed.

### Step 3 – Calibration of HPV transmission and cervical cancer progression models

#### Cluster-level HPV transmission model

The HPV transmission model RHEA was calibrated separately for each cluster of LMICs to the cluster means of indicators of demography (from UN), sexual behavior (from DHS), and HPV prevalence (from literature search, see Section 1 of Supplementary material A for details) of the 132 included LMICs. HPV prevalence data were combined as means in four groups of high-risk types (16, 18, 31/33/45/52/58, and other high-risk types), and calibration was done separately for each group. For each study with HPV prevalence data available, up to seven data-points were included, one for each age-group (15-19, 20-24, 25-29, 30-34, 35-39, 40-49, 50-69). For each cluster, we selected through calibration 100 sets of values of parameters that cannot be directly estimated from data, notably, on under/over-reporting of sexual contact rate, sexual mixing by sexual activity groups, and HPV infection transmission probability. See Section 2 of Supplementary material A for more details of the calibration of the HPV transmission model. See full description of the HPV transmission model RHEA as reported in the appendices of (14) and on the METHIS website (15).

#### Country-level cervical cancer progression model

The calibrated RHEA model outputs together with a second model from the METHIS modelling platform – the cervical cancer progression model, ATLAS - were used to estimate the country-specific progress of cervical cancer elimination provided that country-specific cervical cancer incidence data are available. ATLAS estimates the lifetime number of cervical cancer cases and deaths in a cohort by modifying baseline incidence according to the expected decline in HPV infection. This decline is weighted using HPV type–specific attributable fractions for cervical cancer (20), and accounts for competing mortality. We extracted the baseline age-specific cervical cancer incidence and mortality data from the Global Cancer Observatory (18). See full description of the cervical cancer progression model ATLAS as reported in the appendices of (6) and on the METHIS website (15).

All analyses were performed on R version 4.1.2, OS: linux-gnu, RStudio server 2021.09.01 built 372.

## Data Availability

Population demographic data used are publicly available and can be found at https://population.un.org/wpp/. Sexual behavior data from the DHS program used are publicly available and can be found at https://dhsprogram.com/. HIV-related data from UNAIDS used are publicly available and can be found at https://aidsinfo.unaids.org/. Cervical cancer incidence data used from GLOBOCAN publicly available and can be found at https://gco.iarc.fr/en. HPV prevalence data used identified from literature search are publicly available. The list of articles included can be found in eTable 10.

## Acknowledgements

We acknowledge Feixue Wei and Lucas Dufour for their contribution in collecting HPV prevalence data.

## Disclaimer

Where authors are identified as personnel of the International Agency for Research on Cancer/World Health Organization, the authors alone are responsible for the views expressed in this article and they do not necessarily represent the decisions, policy or views of the International Agency for Research on Cancer /World Health Organization. The designations used and the presentation of the material in this Article do not imply the expression of any opinion whatsoever on the part of WHO and the IARC about the legal status of any country, territory, city, or area, or of its authorities, or concerning the delimitation of its frontiers or boundaries.

## Contributions

IM and IB were responsible for conceptualization, funding acquisition, resources and supervision. IM was responsible for project administration. AM, ME and IA were responsible for data curation. IM, AM, ME and DG were responsible for the formal analysis. IM, AM, ME, AG, DG and IB were responsible for investigation and methodology. DG was responsible for software. AG and DG were responsible for validation. IM, AM and IB were responsible for visualization, and writing – original draft preparation. All co-authors were responsible for writing – review & editing.

