## Supplementary material A for "A unified modeling platform for informing cervical cancer prevention policy decisions in 132 low- and middle-income countries"

### 1 HPV prevalence literature search

For the calibration of our HPV transmission model (RHEA), age- and type-specific prevalence of high-risk HPV types was the main calibration target. In the calibration process, the prevalence for the whole model population was required to reproduce closely the observed prevalence at the country level. The observed HPV prevalence should ideally stem from studies of the general population in a country obtained through random sampling. However, many studies are not conducted in such condition. In such a case, restricting to the part of the study population with normal cytological could still provide suitable data. The exact study data selection algorithm is depicted by the following Figure.

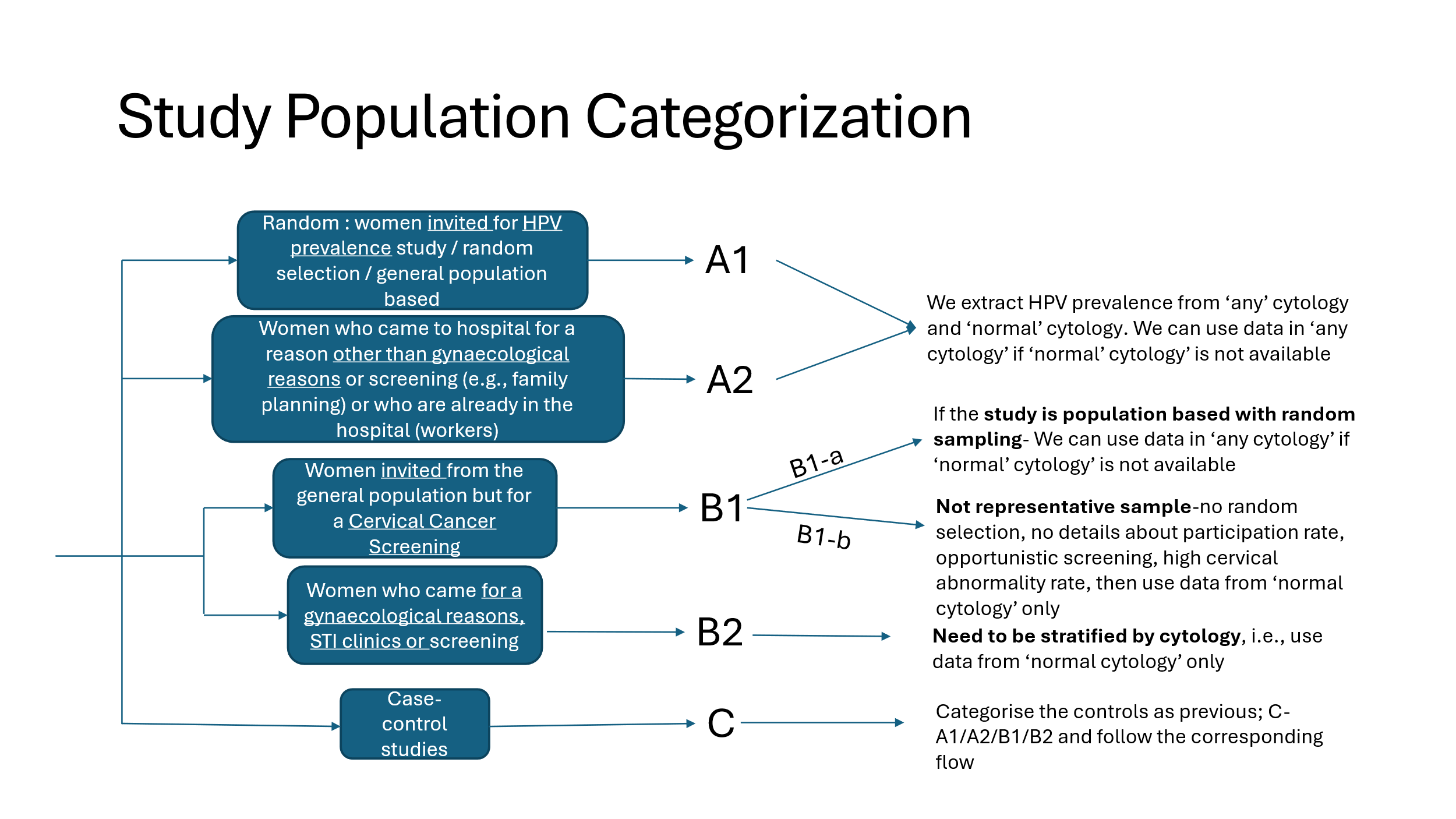

Moreover, the following exclusion criterions were defined to identify suitable studies:

- Studies with sample size of less than 100 women
- Studies not reporting HPV prevalence
- Studies only among high-risk population such as women living with HIV and sex-workers
- Studies only among women with cervical cancer or cervical lesions
- Studies only among pregnant women
- Studies with same baseline population (only one of them with latest and complete HPV prevalence data was included)
- Studies only among HPV positive women

To search for HPV prevalence studies of the 132 low- and middle-income countries (LMICs) listed in eTable 1 from literature we relied on two previous comprehensive systematic reviews, Bruni et al. (1) and Wei et al. (2), and an additional complementary PubMed search.

- **Bruni et al.:** This HPV prevalence systematic review by IARC/ICO HPV Information Centre from 2010 included data among women with normal cytology worldwide, hence suitable data for our modelling purpose. Using the online country report on the IARC/ICO HPV Information Centre, we retrieved all included HPV prevalence studies.
- **Wei et al.:** This HPV prevalence systematic review by us at IARC from 2024 while focusing on invasive cancer also included in the search women with normal cytology. All useful studies for modelling were identified during the search.
- **Additional complementary PubMed search**: As the review by Bruni et al. already identified several studies for most of the countries, we only conducted additional search for countries for which no studies at all had yet been identified. We conducted a search in PubMed on 15-01-2024. The search strategy was “*Papillomavirus infections [MeSH Terms] OR HPV [Text Word] OR papillomavirus*[Text Word] OR papillomaviridae [MeSH Terms] AND (Country Name [Text Word]).*”

See eTable 10 for the list of included HPV prevalence studies. For all identified eligible studies, we extracted the available age- and type-specific HPV prevalence data.

### 2 HPV transmission Model Parametrization and Calibration procedure

The full description of the structure and mathematical formulation of the HPV transmission model RHEA has previously been reported in the appendices of Man et al. (3).

The model calibration procedure was as follows. The model was calibrated to each of the 7 clusters of LMICs independently. The parameter values regarding the type-specific progression, clearance rates and waning rates of natural immunity were independent of the cluster and were fixed to those estimated for the extensively validated cervical cancer progression model (4, 5) and as indicated in eTable 9. Sexual behaviour parameters directly estimable through data included age-specific sexual contact rates (eFigure 8) and sexual contact mixing patterns (eFigure 9). See eTable 9 for the values and references of all fixed parameters.

Model parameters not estimable through data were calibrated to fit the observed HPV prevalence obtained through the literature review described above. The observed HPV prevalence target was adjusted for the average proportion of sexually active women by cluster, computed based on DHS data. This adjustment is needed due to the fact that HPV prevalence studies are usually done in sexually active women, whereas the model outputs HPV prevalence estimates for the general female population. HPV prevalence data were combined as means in four groups of high-risk types (16, 18, 31/33/45/52/58, and other high-risk types), and calibration was done separately for each group.

The parameter values obtained in this calibration step consist of four type-specific transmission probabilities, four assortativeness adjustment parameters, and two adjustment parameters to account for under/over-reporting of sexual contact rate in the DHS (one adjustment parameter for ages 10-19, and one for ages 20-79). See eTable 9 for the ranges of the prior distribution used for the calibrated parameters. Uniform prior distributions were used. Hundred best-fitting parameter sets were obtained through an Approximate Bayesian Computation scheme with the Lenormand method using R package EasyABC (6). For some parameters a two-step fine tuning of the calibration was performed, by restricting the range of the uniform prior distributions to the inter-quartile range of the first calibration attempt. Log-likelihood of the observed HPV prevalence data given the simulated HPV prevalence under a binomial distribution was used as target statistics with an acceptance threshold of 0.05. For each parameter set, log-likelihood was computed at year 150 of simulation after the burn-in period after the model prevalence has stabilized. See Figure 2 from the main text for the fit to the HPV target prevalence and eFigure 10 for the posterior distribution of the calibrated parameters.

### 3 Supplementary tables

**eTable 1. Overview of the 132 low- and middle-income countries included and their data availability.**
a. Income group from World Bank classification of year 2025(7).
b. Sexual behaviour data from the Demographic and Health Surveys according to DHS STATcompiler (8); the percentage corresponds to the proportion of sexual behaviour indicators given in eTable 2 available; for each indicator we took the most value in the most recent survey. * Country aggregated data available when extracted using DHS STATcompiler for the clustering step. but no individual data available, which was needed for the model calibration step.
c. HPV prevalence data from literature search (see Section 1 for the description of the search).
d. Cervical cancer incidence data from GLOBOCAN 2022 (9); coding of quality level used by GLOBOCAN (see coding dictionary below the table).

| **Country** | **Income group^a^** | **Sexual behaviour^b^** | **HPV prevalence^c^** | **Cervical cancer incidence^d^** |
| --- | --- | --- | --- | --- |
| Afghanistan | Low income | >50% DHS data | No data | 9 |
| Albania | Upper middle income | >50% DHS data | Yes | 2a |
| Algeria | Upper middle income | No data | Yes | 2b |
| Angola | Lower middle income | >50% DHS data | No data | 2a |
| Argentina | Upper middle income | No data | Yes | 3c |
| Armenia | Upper middle income | >50% DHS data | Yes | 3c |
| Azerbaijan | Upper middle income | >50% DHS data | No data | 3c |
| Bangladesh | Lower middle income | >50% DHS data | Yes | 9 |
| Belarus | Upper middle income | No data | No data | 1 |
| Belize | Upper middle income | No data | No data | 3c |
| Benin | Lower middle income | >50% DHS data | No data | 2b |
| Bhutan | Lower middle income | No data | Yes | 9 |
| Bolivia | Lower middle income | >50% DHS data | No data | 3c |
| Bosnia and Herzegovina | Upper middle income | No data | No data | 3a |
| Botswana | Upper middle income | <50% DHS data * | Yes | 1 |
| Brazil | Upper middle income | >50% DHS data | Yes | 3c |
| Burkina Faso | Low income | >50% DHS data | No data | 4 |
| Burundi | Low income | >50% DHS data | No data | 9 |
| Cabo Verde | Lower middle income | No data | No data | 1 |
| Cambodia | Lower middle income | >50% DHS data | No data | 9 |
| Cameroon | Lower middle income | >50% DHS data | No data | 9 |
| Central African Republic | Low income | >50% DHS data | No data | 9 |
| Chad | Low income | >50% DHS data | Yes | 2a |
| China | Upper middle income | No data | No data | 2b |
| Colombia | Upper middle income | >50% DHS data | Yes | 3c |
| Comoros | Lower middle income | >50% DHS data | No data | 9 |
| Congo, Dem. Rep. | Low income | >50% DHS data | No data | 9 |
| Congo, Rep. | Lower middle income | >50% DHS data | No data | 2a |
| Costa Rica | Upper middle income in 2023, after high income | No data | Yes | 1 |
| Côte d’Ivoire | Lower middle income | >50% DHS data | No data | 2a |
| Cuba | Upper middle income | No data | No data | 3c |
| Djibouti | Lower middle income | No data | No data | 9 |
| Dominica | Upper middle income | No data | No data | No data |
| Dominican Republic | Upper middle income | >50% DHS data | No data | 3c |
| Ecuador | Upper middle income | <50% DHS data | No data | 3c |
| Egypt, Arab Rep. | Lower middle income | <50% DHS data | No data | 2a |
| El Salvador | Upper middle income | <50% DHS data | No data | 3c |
| Equatorial Guinea | Upper middle income | No data | No data | 9 |
| Eritrea | Low income | >50% DHS data * | No data | 9 |
| Eswatini | Lower middle income | >50% DHS data | Yes | 1 |
| Ethiopia | Low income in 2023, after unclassified | >50% DHS data | Yes | 2a |
| Fiji | Upper middle income | No data | Yes | 3c |
| Gabon | Upper middle income | >50% DHS data | No data | 1 |
| Gambia, The | Low income | >50% DHS data | No data | 1 |
| Georgia | Upper middle income | No data | Yes | 3c |
| Ghana | Lower middle income | >50% DHS data | Yes | 2a |
| Grenada | Upper middle income | No data | No data | No data |
| Guatemala | Upper middle income | >50% DHS data | No data | 3c |
| Guinea | Lower middle income | >50% DHS data | Yes | 2a |
| Guinea-Bissau | Low income | No data | No data | 9 |
| Guyana | Upper middle income in 2021, after high income | >50% DHS data | No data | 3c |
| Haiti | Lower middle income | >50% DHS data | No data | 3c |
| Honduras | Lower middle income | >50% DHS data | Yes | 2a |
| India | Lower middle income | >50% DHS data | Yes | 2b |
| Indonesia | Upper middle income | >50% DHS data | No data | 9 |
| Iran, Islamic Rep. | Upper middle income | No data | Yes | 2b |
| Iraq | Upper middle income | No data | No data | 1 |
| Jamaica | Upper middle income | No data | Yes | 3c |
| Jordan | Lower middle income | <50% DHS data | No data | 1 |
| Kazakhstan | Upper middle income | >50% DHS data | No data | 1 |
| Kenya | Lower middle income | >50% DHS data | No data | 2b |
| Kiribati | Lower middle income | No data | No data | No data |
| Korea, Dem. People's Rep. | Low income | No data | No data | 9 |
| Kosovo | Upper middle income | No data | No data | No data |
| Kyrgyz Republic | Lower middle income | >50% DHS data | No data | 3c |
| Lao PDR | Lower middle income | No data | Yes | 9 |
| Lebanon | Lower middle income | No data | No data | 1 |
| Lesotho | Lower middle income | >50% DHS data | No data | 9 |
| Liberia | Low income | >50% DHS data | No data | 9 |
| Libya | Upper middle income | No data | No data | 2a |
| Madagascar | Low income | >50% DHS data | No data | 9 |
| Malawi | Low income | >50% DHS data | No data | 2a |
| Malaysia | Upper middle income | No data | No data | 9 |
| Maldives | Upper middle income | >50% DHS data | No data | 9 |
| Mali | Low income | >50% DHS data | No data | 2a |
| Marshall Islands | Upper middle income | No data | No data | No data |
| Mauritania | Lower middle income | >50% DHS data | No data | 9 |
| Mauritius | Upper middle income | No data | No data | 1 |
| Mexico | Upper middle income | <50% DHS data | Yes | 3c |
| Micronesia, Fed. Sts. | Lower middle income | No data | No data | No data |
| Moldova | Upper middle income | >50% DHS data | No data | 3b |
| Mongolia | Upper middle income | No data | Yes | 1 |
| Montenegro | Upper middle income | No data | No data | 3b |
| Morocco | Lower middle income | <50% DHS data | No data | 2b |
| Mozambique | Low income | >50% DHS data | Yes | 2b |
| Myanmar | Lower middle income | >50% DHS data | No data | 9 |
| Namibia | Upper middle income | >50% DHS data | No data | 1 |
| Nepal | Lower middle income | >50% DHS data | Yes | 9 |
| Nicaragua | Lower middle income | >50% DHS data | No data | 3c |
| Niger | Low income | >50% DHS data | No data | 2a |
| Nigeria | Lower middle income | >50% DHS data | Yes | 2b |
| North Macedonia | Upper middle income | No data | No data | 3b |
| Pakistan | Lower middle income | >50% DHS data | Yes | 2a |
| Papua New Guinea | Lower middle income | >50% DHS data | No data | 9 |
| Paraguay | Upper middle income | <50% DHS data | No data | 3c |
| Peru | Upper middle income | >50% DHS data | No data | 3c |
| Philippines | Lower middle income | >50% DHS data | No data | 2b |
| Rwanda | Low income | >50% DHS data | Yes | 2a |
| Samoa | Lower middle income | No data | No data | 9 |
| São Tomé and Príncipe | Lower middle income | >50% DHS data | No data | 9 |
| Senegal | Lower middle income | >50% DHS data | Yes | 9 |
| Serbia | Upper middle income | No data | No data | 3b |
| Sierra Leone | Low income | >50% DHS data | No data | 9 |
| Solomon Islands | Lower middle income | No data | No data | 9 |
| Somalia | Low income | No data | No data | 9 |
| South Africa | Upper middle income | >50% DHS data | Yes | 2a |
| South Sudan | Low income | No data | No data | 9 |
| Sri Lanka | Lower middle income | <50% DHS data | No data | 2a |
| St. Lucia | Upper middle income | No data | No data | 3c |
| St. Vincent and the Grenadines | Upper middle income | No data | Yes | No data |
| Sudan | Low income | <50% DHS data | No data | 2a |
| Suriname | Upper middle income | No data | No data | 3c |
| Syrian Arab Republic | Low income | No data | No data | 3c |
| Tajikistan | Lower middle income | >50% DHS data | No data | 3c |
| Tanzania | Lower middle income | >50% DHS data | Yes | 2b |
| Thailand | Upper middle income | <50% DHS data | Yes | 2b |
| Timor-Leste | Lower middle income | >50% DHS data | No data | 9 |
| Togo | Low income | >50% DHS data | No data | 9 |
| Tonga | Upper middle income | No data | No data | No data |
| Trinidad and Tobago | Upper middle income in 2005, after high income | >50% DHS data | No data | 3c |
| Tunisia | Lower middle income | <50% DHS data | Yes | 2b |
| Türkiye | Upper middle income | <50% DHS data | Yes | 2b |
| Turkmenistan | Upper middle income | <50% DHS * | No data | 9 |
| Tuvalu | Upper middle income | No data | No data | No data |
| Uganda | Low income | >50% DHS data | Yes | 2b |
| Ukraine | Upper middle income | >50% DHS data | No data | 1 |
| Uzbekistan | Lower middle income | >50% DHS data | No data | 3c |
| Vanuatu | Lower middle income | No data | Yes | 9 |
| Venezuela, RB | Upper middle income in 2019, after unclassified | No data | No data | 3c |
| Vietnam | Lower middle income | >50% DHS data | Yes | 2b |
| West Bank and Gaza | Lower middle income | No data | No data | 9 |
| Yemen, Rep. | Low income | <50% DHS data | No data | 1 |
| Zambia | Lower middle income | >50% DHS data | No data | 2a |
| Zimbabwe | Lower middle income | >50% DHS data | No data | 2b |

GLOBOCAN cervical cancer incidence quality level coding
1 = National (or sub-national with coverage greater than 50%) rates projected to 2022;
2a = Most recent rates from a single registry applied to 2022 population
2b = Weighted/simple average of the most recent sub-national rates applied to 2022 population
3a = Estimated from national mortality estimates by modelling, using mortality:incidence ratios derived from country-specific cancer registry data
3b = Estimated from national mortality estimates by modelling, using mortality:incidence ratios derived from cancer registry data in neighbouring countries
3c = Estimated from national mortality estimates by modelling, using mortality:incidence ratios derived from survival estimation
4 ="All sites" estimates from neighbouring countries partitioned using frequency data
9 = No data: the rates are those of neighbouring countries or registries in the same area

**eTable *2*. Overview of included sexual behaviour indicators from the Demographic and Health Surveys.**
Sexual behaviour indicators from the Demographic and Health Surveys (DHS) included in the clustering step. We took the most value in the most recent survey from DHS STATcompiler (8).

| **Indicator** | **Definition** |
| --- | --- |
| Women giving birth by age 15 | Percentage of women who gave birth by age 15 |
| Women giving birth by age 18 | Percentage of women who gave birth by age 18 |
| Women giving birth by age 20 | Percentage of women who gave birth by age 20 |
| Women giving birth by age 22 | Percentage of women who gave birth by age 22 |
| Women giving birth by age 25 | Percentage of women who gave birth by age 25 |
| Number of wives: Two or more wives | Percentage of currently married or in union men who have two or more wives |
| Women first married by exact age 15 | Percentage of women who were first married by exact age 15 |
| Women first married by exact age 18 | Percentage of women who were first married by exact age 18 |
| Women first married by exact age 20 | Percentage of women who were first married by exact age 20 |
| Women first married by exact age 22 | Percentage of women who were first married by exact age 22 |
| Women first married by exact age 25 | Percentage of women who were first married by exact age 25 |
| Women never married | Percentage of women who were never married |
| Men first married by exact age 15 | Percentage of men who were first married by exact age 15 |
| Men first married by exact age 18 | Percentage of men who were first married by exact age 18 |
| Men first married by exact age 20 | Percentage of men who were first married by exact age 20 |
| Men first married by exact age 22 | Percentage of men who were first married by exact age 22 |
| Men first married by exact age 25 | Percentage of men who were first married by exact age 25 |
| Men never married | Percentage of men who were never married |
| Age difference between man and woman is 10+ years | Percentage of currently married women age 15-24 for whom the age difference with her partner is 10+ years |
| Age difference between man and woman is 5-9 years | Percentage of currently married women age 15-24 for whom the age difference with her partner is 5-9 years |
| Age difference between man and woman is <5 years | Percentage of currently married women age 15-24 for whom the age difference with her partner is <5 years |
| First sexual intercourse by exact age 15 [Women] | Percentage of women who had first sexual intercourse by exact age 15 |
| First sexual intercourse by exact age 18 [Women] | Percentage of women who had first sexual intercourse by exact age 18 |
| First sexual intercourse by exact age 20 [Women] | Percentage of women who had first sexual intercourse by exact age 20 |
| First sexual intercourse by exact age 22 [Women] | Percentage of women who had first sexual intercourse by exact age 22 |
| First sexual intercourse by exact age 25 [Women] | Percentage of women who had first sexual intercourse by exact age 25 |
| Never had sexual intercourse [Women] | Percentage of women who never had intercourse |
| First sexual intercourse by exact age 15 [Men] | Percentage of men who had first sexual intercourse by exact age 15 |
| First sexual intercourse by exact age 18 [Men] | Percentage of men who had first sexual intercourse by exact age 18 |
| First sexual intercourse by exact age 20 [Men] | Percentage of men who had first sexual intercourse by exact age 20 |
| First sexual intercourse by exact age 22 [Men] | Percentage of men who had first sexual intercourse by exact age 22 |
| First sexual intercourse by exact age 25 [Men] | Percentage of men who had first sexual intercourse by exact age 25 |
| Never had sexual intercourse [Men] | Percentage of men who never had intercourse |
| Higher-risk Sex [Women] | Percentage of women aged 15-49 who have had sexual intercourse with more than one partner in the last 12 months. |
| Condom use during higher-risk sex [Women] | Percentage of women aged 15-49 who had more than one partner in the past 12 months reporting the use of a condom during their last sexual intercourse. |
| Mean number of sexual partners in lifetime [Women] | Mean number of sexual partners in her lifetime among women who ever had sexual intercourse |
| Higher-risk Sex [Men] | Percentage of men aged 15-49 who have had sexual intercourse with more than one partner in the last 12 months. |
| Condom use during higher-risk sex [Men] | Percentage of men aged 15-49 who had more than one partner in the past 12 months reporting the use of a condom during their last sexual intercourse. |
| Mean number of sexual partners in lifetime [Men] | Mean number of sexual partners in his lifetime among men who ever had sexual intercourse |
| Cumulative prevalence of concurrent sexual partners [Women] | Percentage of women who had two (or more) sexual partners that were concurrent anytime during the 12 months preceding the survey |
| Cumulative prevalence of concurrent sexual partners [Men] | Percentage of men who had two (or more) sexual partners that were concurrent anytime during the 12 months preceding the survey |
| Ever paid for sexual intercourse [Men] | Percentage of men who ever paid for sexual intercourse |
| Paid for sexual intercourse in the past 12 months [Men] | Percentage of men who paid for sexual intercourse in the 12 months preceding the survey |
| Age-mixing in sexual partnerships [Young women] | Percentage of young women 15-19 who have had sex in the preceding 12 months with a partner who is 10 or more years older than themselves |
| Age-mixing in sexual partnerships [Young men] | Percentage of young men 15-19 who have had sex in the preceding 12 months with a partner who is 10 or more years older than themselves |

**eTable 3. Membership probability by country under the 4-clustering.**
Membership probabilities representing the likelihood to belong to given cluster estimated with the Gaussian Mixture Model used for the clustering step. Clusters A-D are ordered from top to bottom in Figure 1 of the main text. Dark shades of green indicate high membership probabilities.

| **Country** | **Membership probability** | | | |
| --- | --- | --- | --- | --- |
|  | **Cluster A** | **Cluster B** | **Cluster C** | **Cluster D** |
|  | (southern Africa) | (central Africa and America) | (southeastern and central Asia and Europe) | (northern Africa and south Asia) |
| Afghanistan | 0 | 0.000000141 | 0.0000452 | 0.999955 |
| Albania | 0 | 0.000000094 | 0.993828 | 0.006172 |
| Angola | 0 | 1 | 2.16E-08 | 1.28E-09 |
| Armenia | 2.48E-279 | 0.00000113 | 0.999998 | 0.000000921 |
| Azerbaijan | 0 | 3.58E-08 | 1 | 0.000000439 |
| Bangladesh | 0 | 4.79E-08 | 5.26E-24 | 1 |
| Benin | 0 | 0.999151 | 4.12E-08 | 0.000849 |
| Bolivia | 0 | 0.98833 | 0.01167 | 0.000000128 |
| Brazil | 0 | 0.816569 | 0.18343 | 4.39E-08 |
| Burkina Faso | 0 | 0.097488 | 1.48E-11 | 0.902512 |
| Burundi | 0 | 0.0000431 | 0.889175 | 0.110782 |
| Cambodia | 0 | 0.00000335 | 0.998389 | 0.001608 |
| Cameroon | 0 | 1 | 4.18E-15 | 7.66E-08 |
| Central African Republic | 0 | 0.999103 | 1.52E-10 | 0.000897 |
| Chad | 0 | 0.0000485 | 4.7E-18 | 0.999951 |
| Colombia | 0 | 1 | 0.000000279 | 1.66E-13 |
| Comoros | 0 | 0.00000189 | 0.999561 | 0.000437 |
| Congo | 0 | 1 | 7.6E-15 | 2.34E-13 |
| Congo Democratic Republic | 0 | 1 | 3.89E-09 | 0.000000232 |
| Cote d'Ivoire | 0 | 0.999984 | 1.21E-12 | 0.0000163 |
| Dominican Republic | 0 | 0.999999 | 0.00000081 | 1.26E-10 |
| Eritrea | 0 | 8.74E-11 | 3.27E-09 | 1 |
| Eswatini | 0.975217 | 0.024783 | 1.33E-16 | 4.31E-28 |
| Ethiopia | 0 | 0.000451 | 1.15E-10 | 0.999549 |
| Gabon | 0 | 1 | 3.51E-22 | 7.47E-20 |
| Gambia | 0 | 1.59E-10 | 2.7E-09 | 1 |
| Ghana | 0 | 0.986675 | 0.005295 | 0.00803 |
| Guatemala | 0 | 0.99712 | 0.00288 | 0.00000018 |
| Guinea | 0 | 0.00000964 | 3.81E-14 | 0.99999 |
| Haiti | 0 | 1 | 2.04E-09 | 1.45E-14 |
| Honduras | 0 | 0.999795 | 0.000205 | 6.75E-09 |
| India | 0 | 5.94E-09 | 0.00019 | 0.99981 |
| Indonesia | 0 | 0.01661 | 0.919393 | 0.063997 |
| Kazakhstan | 5.27E-56 | 0.000112 | 0.999887 | 0.0000014 |
| Kenya | 0 | 0.999993 | 0.00000584 | 0.00000123 |
| Kyrgyz Republic | 6.19E-82 | 0.00000173 | 0.999998 | 9.97E-11 |
| Lesotho | 1 | 1.02E-08 | 1.08E-17 | 7.28E-27 |
| Liberia | 0 | 1 | 9.71E-18 | 8.24E-16 |
| Madagascar | 0 | 0.9997 | 0.000257 | 0.0000425 |
| Malawi | 0 | 0.999887 | 6.88E-08 | 0.000113 |
| Maldives | 2.22E-138 | 0.00000123 | 0.828166 | 0.171833 |
| Mali | 0 | 0.00000179 | 4.63E-18 | 0.999998 |
| Mauritania | 0 | 5.02E-17 | 9.19E-11 | 1 |
| Moldova | 1.52E-180 | 0.001192 | 0.998808 | 0.000000819 |
| Mozambique | 0 | 1 | 2.65E-12 | 1.82E-09 |
| Myanmar | 0 | 2.54E-16 | 0.98368 | 0.01632 |
| Namibia | 1 | 4.94E-16 | 1.54E-33 | 2.67E-43 |
| Nepal | 0 | 0.959477 | 0.001455 | 0.039068 |
| Nicaragua | 0 | 0.999989 | 0.0000105 | 7.06E-10 |
| Niger | 0 | 1.25E-09 | 1.16E-26 | 1 |
| Nigeria | 0 | 0.0000149 | 1.16E-11 | 0.999985 |
| Pakistan | 0 | 4.88E-21 | 0.001326 | 0.998674 |
| Papua New Guinea | 0 | 0.006244 | 0.976846 | 0.016909 |
| Peru | 0 | 0.998118 | 0.001882 | 0.000000011 |
| Philippines | 0 | 6.91E-09 | 0.99951 | 0.00049 |
| Rwanda | 0 | 0.0000234 | 0.998608 | 0.001368 |
| Sao Tome and Principe | 0 | 0.999949 | 0.0000287 | 0.000022 |
| Senegal | 0 | 1.87E-15 | 0.00000132 | 0.999999 |
| Sierra Leone | 0 | 0.999992 | 4.48E-13 | 0.00000802 |
| South Africa | 1 | 1.51E-17 | 9.95E-43 | 1.01E-57 |
| Tajikistan | 1.62E-66 | 0.025376 | 0.97404 | 0.000584 |
| Tanzania | 0 | 0.999999 | 0.000000145 | 0.000000772 |
| Timor-Leste | 0 | 1.86E-11 | 0.981237 | 0.018763 |
| Togo | 0 | 0.99765 | 0.0000254 | 0.002324 |
| Uganda | 0 | 0.999951 | 1.87E-09 | 0.0000487 |
| Ukraine | 1.21E-136 | 0.010764 | 0.989235 | 0.000000384 |
| Uzbekistan | 8.98E-82 | 0.012161 | 0.96787 | 0.01997 |
| Vietnam | 3.15E-117 | 0.108001 | 0.89181 | 0.00019 |
| Zambia | 0 | 1 | 5.37E-09 | 2.44E-08 |
| Zimbabwe | 0 | 0.999963 | 0.0000256 | 0.0000117 |

**eTable 4. Cluster mean of sexual behavior indicators of the four main clusters.**
Clusters A-D are ordered from top to bottom in Figure 1 of the main text. Sexual behaviour data from the Demographic and Health Surveys (8).

| **Indictor** | **Cluster mean** | | | |
| --- | --- | --- | --- | --- |
|  | **Cluster A** | **Cluster B** | **Cluster C** | **Cluster D** |
|  | (southern Africa) | (central Africa and America) | (southeastern and central Asia and Europe) | (northern Africa and south Asia) |
| First sexual intercourse by age 15 (women) | 7.230382 | 16.03073 | 2.701093 | 18.74268 |
| First sexual intercourse by age 18 (women) | 43.88773 | 56.8507 | 19.1199 | 54.04009 |
| First sexual intercourse by age 20 (women) | 68.93461 | 76.6924 | 43.19262 | 70.22995 |
| First sexual intercourse by age 22 (women) | 82.23239 | 86.09787 | 63.14372 | 80.08225 |
| First sexual intercourse by age 25 (women) | 89.14386 | 91.97306 | 79.24055 | 87.7459 |
| Never had sex (women) | 1.049698 | 1.524337 | 6.11393 | 3.430138 |
| First married by age 15 (women) | 2.668813 | 9.772918 | 2.847672 | 18.02301 |
| First married by age 18 (women) | 14.3831 | 35.0017 | 17.21183 | 49.32882 |
| First married by age 20 (women) | 26.27465 | 52.99808 | 39.29542 | 65.27278 |
| First married by age 22 (women) | 36.21891 | 66.18113 | 59.20777 | 76.27549 |
| First married by age 25 (women) | 48.78169 | 78.62108 | 77.32326 | 86.24004 |
| Never married (women) | 30.2171 | 8.564814 | 8.209364 | 4.687824 |
| Gave birth by age 15 (women) | 2.554326 | 4.615566 | 0.972656 | 5.802919 |
| Gave birth by age 18 (women) | 20.49577 | 26.86934 | 7.596821 | 29.5627 |
| Gave birth by age 20 (women) | 42.40221 | 48.45356 | 23.63021 | 48.71441 |
| Gave birth by age 22 (women) | 61.24306 | 65.44774 | 44.96866 | 64.57417 |
| Gave birth by age 25 (women) | 78.17264 | 80.23144 | 68.29155 | 79.35294 |
| First sexual intercourse by age 15 (men) | 8.632596 | 15.6139 | 4.860569 | 2.002354 |
| First sexual intercourse by age 18 (men) | 43.33602 | 52.54894 | 23.79974 | 13.30873 |
| First sexual intercourse by age 20 (men) | 67.29879 | 73.47681 | 44.52748 | 28.25461 |
| First sexual intercourse by age 22 (men) | 82.97103 | 85.14148 | 63.56653 | 45.36611 |
| First sexual intercourse by age 25 (men) | 90.7163 | 92.22362 | 79.12253 | 63.04885 |
| Never had sex (men) | 1.694567 | 1.245449 | 4.504151 | 8.045996 |
| First married by age 15 (men) | 0.601207 | 1.082101 | 0.812675 | 0.874083 |
| First married by age 18 (men) | 2.701207 | 7.866124 | 4.311237 | 5.578212 |
| First married by age 20 (men) | 6.824547 | 18.51703 | 11.04747 | 13.44245 |
| First married by age 22 (men) | 14.01851 | 32.27795 | 26.13251 | 25.28158 |
| First married by age 25 (men) | 28.32857 | 53.3733 | 51.70392 | 45.2597 |
| Never married (men) | 37.26197 | 14.23572 | 14.80507 | 14.94907 |
| Age diff >= 10 yrs (marital partners) | 21.01891 | 21.91429 | 12.54658 | 40.39648 |
| Age diff 5-9 yrs (marital partners) | 37.9837 | 34.85282 | 33.28282 | 36.80011 |
| Age diff <5 yrs (marital partners) | 34.78954 | 34.64253 | 43.71667 | 20.5023 |
| Age diff >= 10 yrs (young women) | 6.352716 | 13.04191 | 16.98102 | 38.10597 |
| Age diff >= 10 yrs (young men) | 0.841147 | 1.722197 | 3.467129 | 1.725694 |
| Lifetime number of sexual partners (men) | 9.592958 | 8.566099 | 4.821502 | 4.412978 |
| Concurrent sexual partners (men) | 12.22193 | 15.22091 | 6.918678 | 9.690046 |
| Higher-risk sex (men) | 16.96962 | 18.97993 | 8.072596 | 10.73094 |
| Ever paid for sexual intercourse (men) | 7.365292 | 14.58625 | 10.33202 | 7.07737 |
| Recently paid for sexual intercourse (men) | 1.75996 | 4.784803 | 3.26888 | 2.014359 |
| Two or more wives (men) | 2.885513 | 7.816869 | 4.170045 | 10.94452 |
| Lifetime number of sexual partners (women) | 2.903018 | 2.493406 | 1.548512 | 2.013728 |
| Concurrent sexual partners (women) | 2.559507 | 1.82284 | 1.038328 | 0.915148 |
| Higher-risk sex (women) | 3.737827 | 2.468557 | 1.112422 | 1.171098 |
| Condom use during higher-risk sex (men) | 58.67203 | 29.21989 | 27.5779 | 24.82793 |
| Condom use during higher-risk sex (women) | 64.72797 | 33.73422 | 38.02959 | 23.07761 |
| HIV prevalence (15-19 yrs both sexes) | 19.67565 | 2.715459 | 1.181241 | 0.668902 |
| HIV prevalence (15+ yrs both sexes) | 20.59235 | 2.848194 | 1.154039 | 0.698527 |
| ART coverage (both sexes) | 84.20926 | 68.76549 | 61.23628 | 56.55911 |

**eTable 5. Cluster mean of sexual behavior indicators of the seven region-specific subclusters.**
Clusters I-VII are ordered by decreasing cervical cancer incidence (see eFigure 7). Sexual behaviour data from the Demographic and Health Surveys (8).

| **Indicator** | **Cluster mean** | | | | | | |
| --- | --- | --- | --- | --- | --- | --- | --- |
|  | **Cluster I** | **Cluster II** | **Cluster III** | **Cluster IV** | **Cluster V** | **Cluster VI** | **Cluster VII** |
|  | (southern Africa) | (central Africa) | (northern Africa) | (America) | (southeastern Asia) | (southern Asia) | (central Asia and Europe) |
| First sexual intercourse by age 15 (women) | 7.3 | 18.6 | 20.3 | 11.2 | 4.2 | 15.2 | 0.6 |
| First sexual intercourse by age 18 (women) | 43.9 | 63 | 58 | 44.8 | 22.2 | 45 | 14 |
| First sexual intercourse by age 20 (women) | 69 | 82 | 73.2 | 66.4 | 41.7 | 63.5 | 43.3 |
| First sexual intercourse by age 22 (women) | 82.2 | 90 | 82.1 | 78.3 | 59.2 | 75.6 | 66.6 |
| First sexual intercourse by age 25 (women) | 89.1 | 94 | 88.5 | 87.9 | 75.4 | 86 | 83.5 |
| Never had sex (women) | 1.1 | 0.5 | 3 | 3.4 | 7.6 | 4.6 | 5 |
| First married by age 15 (women) | 2.7 | 10.8 | 18.7 | 7.4 | 4.4 | 17 | 0.7 |
| First married by age 18 (women) | 14.4 | 36.8 | 50.7 | 29.6 | 21 | 47 | 12.5 |
| First married by age 20 (women) | 26.3 | 55.1 | 65.8 | 46.9 | 39.4 | 65 | 39.5 |
| First married by age 22 (women) | 36.3 | 68 | 76.3 | 60.6 | 56.8 | 77 | 62.9 |
| First married by age 25 (women) | 48.8 | 80.1 | 85.7 | 74.2 | 75.1 | 87.9 | 81.4 |
| Never married (women) | 30.2 | 7.7 | 4.7 | 10.9 | 9 | 4.6 | 6.8 |
| Gave birth by age 15 (women) | 2.6 | 5.8 | 6.2 | 2.6 | 1.3 | 4.9 | 0.1 |
| Gave birth by age 18 (women) | 20.6 | 30.1 | 31.1 | 20.8 | 10 | 26.3 | 3.5 |
| Gave birth by age 20 (women) | 42.5 | 52.6 | 50.3 | 40.2 | 24.8 | 45.7 | 20.8 |
| Gave birth by age 22 (women) | 61.3 | 69.4 | 65.7 | 57 | 42.7 | 62.6 | 46.2 |
| Gave birth by age 25 (women) | 78.2 | 83.5 | 79.9 | 72.9 | 64.9 | 78.6 | 71.6 |
| First sexual intercourse by age 15 (men) | 8.6 | 11.1 | 2.3 | 26.3 | 5.6 | 0.9 | 4.5 |
| First sexual intercourse by age 18 (men) | 43.3 | 46.5 | 15.5 | 68.1 | 21.7 | 7.1 | 27.3 |
| First sexual intercourse by age 20 (men) | 67.2 | 70 | 31.9 | 83.7 | 38.4 | 18.4 | 52.9 |
| First sexual intercourse by age 22 (men) | 82.9 | 83.8 | 50.1 | 90.4 | 55.4 | 32.5 | 74.5 |
| First sexual intercourse by age 25 (men) | 90.7 | 91.7 | 66.4 | 94.4 | 71.7 | 54 | 88 |
| Never had sex (men) | 1.7 | 1.2 | 6.6 | 1.3 | 6.5 | 12.1 | 2.9 |
| First married by age 15 (men) | 0.6 | 1 | 0.9 | 1.1 | 0.8 | 0.8 | 0.8 |
| First married by age 18 (men) | 2.7 | 6.9 | 5 | 8.8 | 6.1 | 7.2 | 2.4 |
| First married by age 20 (men) | 6.8 | 16.6 | 11.8 | 20.7 | 14.9 | 18.1 | 7.3 |
| First married by age 22 (men) | 14 | 29.7 | 22.7 | 35.8 | 29.2 | 32.4 | 23.6 |
| First married by age 25 (men) | 28.3 | 50.9 | 41.8 | 56.7 | 52 | 54.4 | 52.5 |
| Never married (men) | 37.2 | 14.1 | 15.7 | 15.4 | 14.9 | 13 | 15 |
| Age diff >= 10 yrs (marital partners) | 21.1 | 24.2 | 49 | 18.5 | 12.5 | 18.3 | 9.5 |
| Age diff 5-9 yrs (marital partners) | 38 | 36.9 | 36.2 | 30.9 | 29.6 | 39 | 36.7 |
| Age diff <5 yrs (marital partners) | 34.8 | 32.3 | 13.6 | 38 | 44.4 | 37.9 | 45.2 |
| Age diff >= 10 yrs (young women) | 6.4 | 14 | 42.9 | 11.4 | 19 | 25.6 | 14.6 |
| Age diff >= 10 yrs (young men) | 1 | 1.2 | 1.4 | 2.7 | 2.5 | 2.4 | 5 |
| Lifetime number of sexual partners (men) | 9.6 | 8.7 | 4 | 8.9 | 3.8 | 5.4 | 6.3 |
| Concurrent sexual partners (men) | 12.2 | 16.7 | 10.4 | 13.5 | 6.2 | 7.7 | 8.3 |
| Higher-risk sex (men) | 17 | 20.5 | 12.3 | 17.6 | 6.2 | 6.2 | 10.1 |
| Ever paid for sexual intercourse (men) | 7.4 | 13.8 | 6.8 | 17.6 | 6.1 | 8 | 15 |
| Recently paid for sexual intercourse (men) | 1.8 | 5.5 | 2.1 | 3.8 | 2.4 | 1.7 | 4.2 |
| Two or more wives (men) | 2.9 | 9.7 | 14.5 | 4.4 | 2.8 | 1.6 | 5 |
| Lifetime number of sexual partners (women) | 2.9 | 2.8 | 1.8 | 2 | 1.6 | 2.4 | 1.5 |
| Concurrent sexual partners (women) | 2.6 | 2.1 | 0.7 | 1.5 | 1.1 | 1.3 | 1.2 |
| Higher-risk sex (women) | 3.7 | 2.9 | 1 | 1.9 | 1.2 | 1.4 | 1.1 |
| Condom use during higher-risk sex (men) | 58.7 | 28.4 | 22.6 | 30.3 | 22.8 | 30.3 | 33 |
| Condom use during higher-risk sex (women) | 64.7 | 27.9 | 20.2 | 44.9 | 30.1 | 30.5 | 48.8 |
| HIV prevalence (15-19 yrs both sexes) | 19.7 | 3.8 | 0.8 | 0.7 | 1 | 0.2 | 1.4 |
| HIV prevalence (15+ yrs both sexes) | 20.7 | 4 | 0.9 | 0.7 | 1 | 0.1 | 1.3 |
| ART coverage (both sexes) | 84.3 | 69.5 | 66.7 | 66.8 | 57.7 | 30 | 56.6 |

**eTable 6. Geography-driven classification and data availability for validation.**
a. Sexual behaviour data from the Demographic and Health Surveys (8); the percentage corresponds to the proportion of sexual behaviour indicators available.

b. Cervical cancer incidence data from GLOBOCAN (10) was only used for validation if DHS data was not available. Hence, NA is indicated when DHS data was available.

| **Country** | **Geography-driven classified cluster** | **DHS data availability ^a^** | **GLOBOCAN data availability ^b^** |
| --- | --- | --- | --- |
| Botswana | I | 1-50% DHS data | NA |
| Egypt, Arab Rep. | III | 1-50% DHS data | NA |
| Morocco | III | 1-50% DHS data | NA |
| Sudan | III | 1-50% DHS data | NA |
| Tunisia | III | 1-50% DHS data | NA |
| Yemen, Rep. | III | 1-50% DHS data | NA |
| Ecuador | IV | 1-50% DHS data | NA |
| El Salvador | IV | 1-50% DHS data | NA |
| Mexico | IV | 1-50% DHS data | NA |
| Paraguay | IV | 1-50% DHS data | NA |
| Sri Lanka | V | 1-50% DHS data | NA |
| Thailand | V | 1-50% DHS data | NA |
| Jordan | VII | 1-50% DHS data | NA |
| Türkiye | VII | 1-50% DHS data | NA |
| Turkmenistan | VII | 1-50% DHS data | NA |
| Equatorial Guinea | II | No data | GLOBOCAN data |
| Mauritius | II | No data | GLOBOCAN data |
| Algeria | III | No data | GLOBOCAN data |
| Cabo Verde | III | No data | GLOBOCAN data |
| Djibouti | III | No data | GLOBOCAN data |
| Guinea-Bissau | III | No data | GLOBOCAN data |
| Somalia | III | No data | GLOBOCAN data |
| South Sudan | III | No data | GLOBOCAN data |
| Argentina | IV | No data | GLOBOCAN data |
| Belize | IV | No data | GLOBOCAN data |
| Costa Rica | IV | No data | GLOBOCAN data |
| Cuba | IV | No data | GLOBOCAN data |
| Jamaica | IV | No data | GLOBOCAN data |
| Suriname | IV | No data | GLOBOCAN data |
| Venezuela, RB | IV | No data | GLOBOCAN data |
| Fiji | V | No data | GLOBOCAN data |
| Lao PDR | V | No data | GLOBOCAN data |
| Malaysia | V | No data | GLOBOCAN data |
| Samoa | V | No data | GLOBOCAN data |
| Solomon Islands | V | No data | GLOBOCAN data |
| St. Lucia | V | No data | GLOBOCAN data |
| Vanuatu | V | No data | GLOBOCAN data |
| Bhutan | VI | No data | GLOBOCAN data |
| Belarus | VII | No data | GLOBOCAN data |
| Bosnia and Herzegovina | VII | No data | GLOBOCAN data |
| Georgia | VII | No data | GLOBOCAN data |
| Iran, Islamic Rep. | VII | No data | GLOBOCAN data |
| Iraq | VII | No data | GLOBOCAN data |
| Lebanon | VII | No data | GLOBOCAN data |
| Libya | VII | No data | GLOBOCAN data |
| Mongolia | VII | No data | GLOBOCAN data |
| Montenegro | VII | No data | GLOBOCAN data |
| North Macedonia | VII | No data | GLOBOCAN data |
| Serbia | VII | No data | GLOBOCAN data |
| Syrian Arab Republic | VII | No data | GLOBOCAN data |
| West Bank and Gaza | VII | No data | GLOBOCAN data |
| Dominica | IV | No data | No data |
| Grenada | IV | No data | No data |
| Kiribati | V | No data | No data |
| Marshall Islands | V | No data | No data |
| Micronesia, Fed. Sts. | V | No data | No data |
| St. Vincent and the Grenadines | V | No data | No data |
| Tonga | V | No data | No data |
| Tuvalu | V | No data | No data |
| Kosovo | VII | No data | No data |
| China | Not classified | NA | NA |
| Korea, Dem. People's Rep. | Not classified | NA | NA |

**eTable 7. Validation by membership probability for countries with partially available Demographic and Health Surveys sexual behaviour data.**
Membership probability estimated using the fitted the GMM. Dark shades of green indicate high membership probabilities. The superscript number indicates the ranking received by the geography-driven classified cluster (also indicate3d in bold) based on the classification based on GMM estimated membership probabilities. Countries with some data but for only <50% indicators were included. We also validated the reassignment of the four countries that were had clustering that did not respect with the geographical separation.

| **Country** | **Geography-driven classified cluster** | **Membership probability** | | | | | | |
| --- | --- | --- | --- | --- | --- | --- | --- | --- |
|  |  | **Cluster I** | **Cluster II** | **Cluster III** | **Cluster IV** | **Cluster V** | **Cluster VI** | **Cluster VII** |
|  |  | (southern Africa) | (central Africa) | (northern Africa) | (America) | (southeastern Asia) | (southern Asia) | (central Asia and Europe) |
| Botswana | I | **0.201991^1^** | 5.69E-10 | 8.07E-51 | 2.91E-59 | 1.27E-52 | 0 | 1.72E-85 |
| Ecuador | IV | 0 | 0.006345 | 0.011549 | **0.083546 ^1^** | 2.82E-04 | 6.49E-64 | 1.14E-24 |
| Egypt | III | 0 | 3.38E-06 | **0.001884 ^2^** | 2.40E-08 | 0.076089 | 0.008593031 | 7.59E-08 |
| El Salvador | IV | 0 | 1.83E-06 | 1.94E-05 | **1.74E-12 ^5^** | 0.004286 | 3.82E-18 | 1.22E-05 |
| Jordan | VII | 0 | 6.58E-09 | 3.71E-05 | 8.67E-04 | 0.01458 | 7.56E-130 | **0.02676 ^2^** |
| Mexico | IV | 0 | 0.007493 | 0.021467 | **0.279875 ^1^** | 8.32E-05 | 8.68E-63 | 4.65E-34 |
| Morocco | III | 0 | 8.68E-06 | **1.44E-04 ^2^** | 6.12E-06 | 0.002242 | 5.64E-157 | 3.99E-13 |
| Paraguay | IV | 0 | 1.94E-04 | 0.002758 | **0.157358 ^1^** | 0.011834 | 1.21E-82 | 2.72E-11 |
| Sri Lanka | V | 0 | 2.76E-07 | 4.94E-04 | 1.50E-08 | **0.02964 ^1^** | 1.52E-103 | 6.20E-07 |
| Sudan | III | 0 | 1.68E-04 | **3.15E-04 ^1^** | 2.96E-39 | 3.57E-17 | 1.88E-57 | 6.95E-153 |
| Thailand | V | 0 | 2.27E-05 | 0.001241 | 3.91E-05 | **0.038608 ^1^** | 3.16E-57 | 5.69E-04 |
| Tunisia | III | 0 | 1.57E-07 | **0.001681 ^2^** | 4.88E-14 | 0.06353 | 3.70E-15 | 1.77E-07 |
| Turkey | VII | 0 | 3.81E-06 | 0.001238 | 1.72E-07 | 0.059932 | 6.35E-31 | **0.052849 ^2^** |
| Turkmenistan | VII | 0 | 1.45E-07 | 2.62E-04 | 3.88E-06 | 0.031503 | 3.97E-62 | **0.093261 ^1^** |
| Yemen | III | 0 | 0.001345 | **0.008777 ^1^** | 4.00E-13 | 2.25E-08 | 5.34E-13 | 1.07E-82 |
| Burundi | II | 0 | 3.80E-04 | 5.49E-04 | 4.79E-13 | 0.002428 | 2.29E-56 | 4.29E-07 |
| Comoros | II | 0 | 3.38E-04 | 0.007776 | 6.93E-11 | 5.14E-04 | 8.39E-64 | 9.07E-34 |
| Nepal | VI | 0 | 0.002414 | 0.001984 | 6.29E-14 | 1.33E-05 | 0.001302773 | 5.51E-32 |
| Rwanda | II | 1.27E-91 | 2.80E-08 | 8.50E-07 | 4.33E-04 | 0.001475 | 1.03E-182 | 0.00227 |

**eTable 8. Validation by comparison with GLOBOCAN cervical cancer age-standardized incidence.**
ASIR = Aged-standardized incidence rate in cases per 100,000 women per year from GLOBOCAN 2022 (9). Dark shades of green indicate small difference between the cluster median cervical cancer ASIR and the country incidence as reported in eFigure 7. The superscript number indicates the ranking received by the geography-driven classified cluster (also indicated in bold) based on the classification based on difference to the cluster median cervical cancer ASIR.

| **Country** | **Geography-driven classified cluster** | **Cervical cancer ASIR** | **Difference to cluster median cervical cancer ASIR** | | | | | | |
| --- | --- | --- | --- | --- | --- | --- | --- | --- | --- |
|  |  |  | **Cluster I** | **Cluster II** | **Cluster III** | **Cluster IV** | **Cluster V** | **Cluster VI** | **Cluster VII** |
|  |  |  | (southern Africa) | (central Africa) | (northern Africa) | (America) | (southeastern Asia) | (southern Asia) | (central Asia and Europe) |
| Algeria | III | 7.96 | 39.0 | 25.1 | **19.3 ^5^** | 11.5 | 7.6 | 3.3 | 6.2 |
| Argentina | IV | 16.78 | 30.2 | 16.2 | 10.5 | **2.7 ^3^** | 1.3 | 5.5 | 2.6 |
| Belarus | VII | 9.25 | 37.7 | 23.8 | 18.0 | 10.3 | 6.3 | 2.0 | **4.9 ^2^** |
| Belize | IV | 17.27 | 29.7 | 15.7 | 10.0 | **2.2 ^2^** | 1.8 | 6.0 | 3.1 |
| Bhutan | VI | 13.56 | 33.4 | 19.5 | 13.7 | 5.9 | 2.0 | **2.3 ^3^** | 0.6 |
| Bosnia and Herzegovina | VII | 12.33 | 34.6 | 20.7 | 14.9 | 7.2 | 3.2 | 1.1 | **1.8 ^2^** |
| Cabo Verde | III | 16.07 | 30.9 | 16.9 | **11.2 ^5^** | 3.4 | 0.6 | 4.8 | 1.9 |
| Costa Rica | IV | 10.6 | 36.4 | 22.4 | 16.6 | **8.9 ^4^** | 4.9 | 0.7 | 3.5 |
| Cuba | IV | 12.83 | 34.1 | 20.2 | 14.4 | **6.7 ^4^** | 2.7 | 1.6 | 1.3 |
| Djibouti | III | 16.53 | 30.4 | 16.5 | **10.7 ^5^** | 3.0 | 1.0 | 5.3 | 2.4 |
| Equatorial Guinea | II | 33.23 | 13.7 | **0.2 ^1^** | 6.0 | 13.7 | 17.7 | 22.0 | 19.1 |
| Fiji | V | 34.97 | 12.0 | 2.0 | 7.7 | 15.5 | **19.5 ^5^** | 23.7 | 20.8 |
| Georgia | VII | 10.35 | 36.6 | 22.7 | 16.9 | 9.2 | 5.2 | 0.9 | **3.8 ^3^** |
| Guinea-Bissau | III | 34.3 | 12.7 | 1.3 | **7.1 ^2^** | 14.8 | 18.8 | 23.0 | 20.2 |
| Iran, Islamic Rep. | VII | 2.53 | 44.4 | 30.5 | 24.7 | 17.0 | 13.0 | 8.8 | **11.6 ^2^** |
| Iraq | VII | 2.18 | 44.8 | 30.8 | 25.1 | 17.3 | 13.3 | 9.1 | **12.0 ^2^** |
| Jamaica | IV | 20.4 | 26.6 | 12.6 | 6.8 | **0.9 ^1^** | 4.9 | 9.1 | 6.3 |
| Lao PDR | V | 11.95 | 35.0 | 21.1 | 15.3 | 7.6 | **3.6 ^3^** | 0.7 | 2.2 |
| Lebanon | VII | 3.59 | 43.4 | 29.4 | 23.6 | 15.9 | 11.9 | 7.7 | **10.6 ^2^** |
| Libya | VII | 8 | 39.0 | 25.0 | 19.2 | 11.5 | 7.5 | 3.3 | **6.1 ^2^** |
| Malaysia | V | 10.33 | 36.6 | 22.7 | 16.9 | 9.2 | **5.2 ^3^** | 0.9 | 3.8 |
| Mauritius | II | 12.9 | 34.1 | **20.1 ^6^** | 14.3 | 6.6 | 2.6 | 1.6 | 1.2 |
| Mongolia | VII | 20.19 | 26.8 | 12.8 | 7.0 | 0.7 | 4.7 | 8.9 | **6.1 ^3^** |
| Montenegro | VII | 11.96 | 35.0 | 21.1 | 15.3 | 7.5 | 3.6 | 0.7 | **2.2 ^2^** |
| North Macedonia | VII | 6.72 | 40.3 | 26.3 | 20.5 | 12.8 | 8.8 | 4.6 | **7.4 ^2^** |
| Samoa | V | 13.29 | 33.7 | 19.7 | 13.9 | 6.2 | **2.2 ^3^** | 2.0 | 0.9 |
| Serbia | VII | 13.35 | 33.6 | 19.7 | 13.9 | 6.2 | 2.2 | 2.1 | **0.8 ^1^** |
| Solomon Islands | V | 26.15 | 20.8 | 6.9 | 1.1 | 6.7 | **10.6 ^4^** | 14.9 | 12.0 |
| Somalia | III | 26.58 | 20.4 | 6.4 | **0.7 ^1^** | 7.1 | 11.1 | 15.3 | 12.4 |
| South Sudan | III | 21.42 | 25.6 | 11.6 | **5.8 ^2^** | 1.9 | 5.9 | 10.1 | 7.3 |
| St. Lucia | V | 15.72 | 31.3 | 17.3 | 11.5 | 3.8 | **0.2 ^1^** | 4.4 | 1.6 |
| Suriname | IV | 24.2 | 22.8 | 8.8 | 3.0 | **4.7 ^2^** | 8.7 | 12.9 | 10.1 |
| Syrian Arab Republic | VII | 2.51 | 44.5 | 30.5 | 24.7 | 17.0 | 13.0 | 8.8 | **11.6 ^2^** |
| Vanuatu | V | 18.15 | 28.8 | 14.9 | 9.1 | 1.4 | **2.6 ^2^** | 6.9 | 4.0 |
| Venezuela, RB | IV | 22.73 | 24.2 | 10.3 | 4.5 | **3.2 ^1^** | 7.2 | 11.5 | 8.6 |
| West Bank and Gaza | VII | 3.12 | 43.9 | 29.9 | 24.1 | 16.4 | 12.4 | **8.2 ^1^** | 11.0 |

**eTable 9. Input parameters used in the HPV transmission model RHEA.**

| **Notation** | **Description** | **Values / ranges** | **Reference** |
| --- | --- | --- | --- |
| *Demography* | | | |
| $m^{W}(a)$ ,$m^{M}(a)$ | Gender- and age-specific mortality | Estimates for the last five recent years 2021-2025 | UN (11) |
| $b$ | Population sex-specific birth rate | Set such that the total population stays constant given $m^{g}(a)$ and 50%-50% distribution of female and male population influx | NA |
| *Sexual contact behaviour* | | | |
| $p^{W}\left( low \right),p^{W}\left( medium \right),p^{W}\left( high \right),$  $p^{M}\left( low \right),p^{M}\left( medium \right),p^{M}\left( high \right)$ | Population distribution of CSA | 0.80, 0.15, 0.05,  0.80, 0.15, 0.05 by assumption | NA |
| $\rho^{W,age}(a,a')$ | Age mixing matrix in women | See eFigure 9 | Survey data on the age difference between sexual partners. (8) |
| $\rho^{M,age}(a,a')$ | Age mixing matrix in men | Derived from the age matrix in women by transposing it and then rescaling rows to unity. | NA |
| $c^{W}(a)$, $c^{M}(a)$ | Partner acquisition rate in women and men | See eFigure 8 | Survey data on the number of sexual partners by age. (8) |
| $\rho^{W,csa}(l, l')$, $\rho^{M,csa}(l, l')$ | CSA mixing matrix in women and men | Identity matrix by assumption | NA |
| $\varepsilon^{W, age}$, $\varepsilon^{M,age}$ | Age assortativeness adjustment parameters | Derived from model calibration with the following range for candidate values with prior Uniform(0.1, 0.5). See eFigure 10 | NA |
| $\varepsilon^{W, csa}$, $\varepsilon^{M,csa}$ | CSA assortativeness adjustment parameters | Derived from model calibration with the following range for candidate values with prior Uniform(0.1, 0.9). See eFigure 10 | NA |
| $\varepsilon^{10-19}$, $\varepsilon^{20-79}$ | Adjustment parameters for under/over-reporting of sexual contact rate, for age groups 10-19 and 20-79 | Derived from model calibration with the following range for candidate values with initial prior Uniform(0.1, 0.9) within the two-step calibration procedure as described in Section 2. See eFigure 10 | NA |
| $\theta$ | Balance parameter | 0.5 by assumption | NA |
| *HPV natural history* | | | |
| $\beta_{16}, \beta_{18},\beta_{nona}, \beta_{other}$ | Transmission probability of type $i$ per sex act | Derived from model calibration with the following range for candidate values with initial prior Uniform(0.1, 0.9).  **See eFigure 10** | NA |
| $\gamma_{16}, \gamma_{18},\gamma_{nona}, \gamma_{other}$ | Clearance rate from CIN0 | 0.824 per year for HPV 16,  0.955 per year for HPV 18,  1.18 per year for HPV 31/33/45/52/58 and remaining HR HPV types | (4, 5) |
| $\eta_{16}, \eta_{18},\eta_{nona}, \eta_{other}$ | Progression rate from CIN0 to CIN1 | 0.676 per year for HPV 16,  0.545 per year for HPV 18,  0.324 per year for HPV 31/33/45/52/58 and remaining HR HPV types | (4, 5) |
| $\delta_{16,1}, \delta_{18,1}$,$\delta_{nona,1}$, $\delta_{other,1}$ | Clearance rate from CIN1 | 0.133 per year for HPV 16,  0.386 per year for HPV 18,  0.481 per year for HPV 31/33/45/52/58 and remaining HR HPV types | (4, 5) |
| $\delta_{16,2}$,$\delta_{18,2},\delta_{nona,2}$,$\delta_{other,2}$ | Clearance rate from regressive CIN2/3 | 2.10 per year for HPV 16,  2.10 per year for HPV 18,  2.10 per year for HPV 31/33/45/52/58 and remaining HR HPV types | (4, 5) |
| $\nu_{16,1}$,$\nu_{18,1}$,$\nu_{nona,1}$, $\nu_{other,1}$ | Progression rate from CIN1 to regressive CIN2/3 | 0.048 per year for HPV 16,  0.00681 per year for HPV 18,  0.0447 per year for HPV 31/33/45/52/58 and remaining HR HPV types | (4, 5) |
| $\nu_{16,2}$,$\nu_{18,2}$,$\nu_{nona,2}$, $\nu_{other,2}$ | Progression rate from CIN1 to non-regressive CIN2/3 | 0.0454 per year for HPV 16,  0.0450 per year for HPV 18,  0.0110 per year for HPV 31/33/45/52/58 and remaining HR HPV types | (4, 5) |
| $\mu_{16}$, $\mu_{18}$,$\mu_{nona}$, $\mu_{other}$ | Rate of waning natural immunity | 0.0407 per year for HPV 16,  0.0287 per year for HPV 18,  0.0320 per year for HPV 31/33/45/52/58 and remaining HR HPV types | (4, 5) |

**eTable 10. List of HPV prevalence studies included in the calibration of the HPV transmission model RHEA.**

Alibegashvili, T., Clifford, G. M., Vaccarella, S., Baidoshvili, A., Gogiashvili, L., Tsagareli, Z., Kureli, I., Snijders, P. J. F., Heideman, D. A. M., van Kemenade, F. J., Meijer, C. J. L. M., Kordzaia, D., and Franceschi, S. (2011). Human papillomavirus infection in women with and without cervical cancer in Tbilisi, Georgia. *Cancer Epidemiology* **35**, 465-470.

Andall-Brereton, G., Brown, E., Slater, S., Holder, Y., Luciani, S., Lewis, M., and Irons, B. (2017). Prevalence of high-risk human papillomavirus among women in two English-speaking Caribbean countries. *Rev Panam Salud Publica* **41**, e41.

Anh, P. T. H., Hieu, N. T., Herrero, R., Vaccarella, S., Smith, J. S., Thuy, N. T., Nga, N. H., Duc, N. B., Ashley, R., Snijders, P. J. F., Meijer, C. J. L. M., Muñoz, N., Parkin, D. M., and Franceschi, S. (2003). Human papillomavirus infection among women in South and North Vietnam. *International Journal of Cancer* **104**, 213-220.

Ardhaoui, M., Letaief, H., Ennaifer, E., Bougatef, S., Lassili, T., Bel Haj Rhouma, R., Fehri, E., Ouerhani, K., Guizani, I., McHela, M., Chahed, K., Chahed, M. K., Boubaker, M. S., and Bouafif Ben Alaya, N. (2022). The Prevalence, Genotype Distribution and Risk Factors of Human Papillomavirus in Tunisia: A National-Based Study. *Viruses* **14**.

Aruhuri, B., Tarivonda, L., Tenet, V., Sinha, R., Snijders, P. J., Clifford, G., Pang, J., McAdam, M., Meijer, C. J., Frazer, I. H., and Franceschi, S. (2012). Prevalence of cervical human papillomavirus (HPV) infection in Vanuatu. *Cancer Prev Res (Phila)* **5**, 746-53.

Aziz (2018). "Human Papillomavirus (HPV) infection in females with normal cervical cytology: Genotyping and Phylogenetic analysis among women in Punjab, Pakistan".

Banura, C., Franceschi, S., Van Doorn, L. J., Arslan, A., Wabwire-Mangen, F., Mbidde, E. K., Quint, W., and Weiderpass, E. (2008). Infection with human papillomavirus and HIV among young women in Kampala, Uganda. *Journal of Infectious Diseases* **197**, 555-562.

Baussano, I., Tenet, V., Baghdasarova, K., Harutyunyan, Z., Vorsters, A., Heideman, D., Bleeker, M., Rüttimann, R., and Sahakyan, G. (2025). HPV burden in Armenia among unvaccinated women: a series of cross-sectional population-based prevalence surveys. *Vaccine* **62**.

Bouassa, R. S. M., Nodjikouambaye, Z. A., Sadjoli, D., Adawaye, C., Péré, H., Veyer, D., Matta, M., Robin, L., Tonen-Wolyec, S., Moussa, A. M., Koyalta, D., and Belec, L. (2019). High prevalence of cervical high-risk human papillomavirus infection mostly covered by Gardasil-9 prophylactic vaccine in adult women living in N’Djamena, Chad. *PLoS ONE* **14**.

Castellsagué, X., Menéndez, C., Loscertales, M. P., Kornegay, J. R., dos Santos, F., Gómez-Olivé, F. X., Lloveras, B., Abarca, N., Vaz, N., Barreto, A., Bosch, F. X., and Alonso, P. (2001). Human papillomavirus genotypes in rural Mozambique. *Lancet* **358**, 1429-30.

Dai, M., Bao, Y. P., Li, N., Clifford, G. M., Vaccarella, S., Snijders, P. J. F., Huang, R. D., Sun, L. X., Meijer, C. J. L. M., Qiao, Y. L., and Franceschi, S. (2006). Human papillomavirus infection in Shanxi Province, People's Republic of China: A population-based study. *British Journal of Cancer* **95**, 96-101.

Datta, P., Bhatla, N., Dar, L., Patro, A. R., Gulati, A., Kriplani, A., and Singh, N. (2010). Prevalence of human papillomavirus infection among young women in North India. *Cancer Epidemiology* **34**, 157-161.

Demir, E. T., Ceyhan, M., Simsek, M., Gunduz, T., Arlier, S., Aytac, R., Aycan, A. E., and Gurbuz, V. (2012). The prevalence of different HPV types in Turkish women with a normal Pap smear. *J Med Virol* **84**, 1242-7.

Dondog, B., Clifford, G. M., Vaccarella, S., Waterboer, T., Unurjargal, D., Avirmed, D., Enkhtuya, S., Kommoss, F., Wentzensen, N., Snijders, P. J. F., Meijer, C. J. L. M., Franceschi, S., and Pawlita, M. (2008). Human papillomavirus infection in Ulaanbaatar, Mongolia: A population-based study. *Cancer Epidemiology Biomarkers and Prevention* **17**, 1731-1738.

Dutta, S., Begum, R., Mazumder Indra, D., Mandal, S. S., Mondal, R., Biswas, J., Dey, B., Panda, C. K., and Basu, P. (2012). Prevalence of human papillomavirus in women without cervical cancer: a population-based study in Eastern India. *Int J Gynecol Pathol* **31**, 178-183.

Ebrahim, S., Mndende, X. K., Kharsany, A. B., Mbulawa, Z. Z., Naranbhai, V., Frohlich, J., Werner, L., Samsunder, N., Karim, Q. A., and Williamson, A. L. (2016). High Burden of Human Papillomavirus (HPV) Infection among Young Women in KwaZulu-Natal, South Africa. *PLoS One* **11**, e0146603.

Figueiredo Alves, R. R., Turchi, M. D., Santos, L. E., Guimarães, E. M., Garcia, M. M., Seixas, M. S., Villa, L. L., Costa, M. C., Moreira, M. A., and Alves Mde, F. (2013). Prevalence, genotype profile and risk factors for multiple human papillomavirus cervical infection in unimmunized female adolescents in Goiânia, Brazil: a community-based study. *BMC Public Health* **13**, 1041.

Filipi, K., Tedeschini, A., Paolini, F., Celicu, S., Morici, S., Kota, M., Bucaj, E., and De Marco, F. (2010). Genital human papillomavirus infection and genotype prevalence among Albanian women: a cross-sectional study. *J Med Virol* **82**, 1192-6.

Foliaki, S., Brewer, N., Pearce, N., Snijders, P. J., Meijer, C. J., Waqatakirewa, L., Clifford, G. M., and Franceschi, S. (2014). Prevalence of HPV infection and other risk factors in a Fijian population. *Infect Agent Cancer* **9**, 14.

Fonseca, A. J., Taeko, D., Chaves, T. A., Da Costa Amorim, L. D., Murari, R. S. W., Miranda, A. E., Chen, Z., Burk, R. D., and Ferreira, L. C. L. (2015). HPV infection and cervical screening in socially isolated indigenous women inhabitants of the amazonian rainforest. *PLoS ONE* **10**.

Franceschi, S., Rajkumar, R., Snijders, P. J. F., Arslan, A., Mahé, C., Plummer, M., Sankaranarayanan, R., Cherian, J., Meijer, C. J. L. M., and Weiderpass, E. (2005). Papillomavirus infection in rural women in southern India. *British Journal of Cancer* **92**, 601-606.

Gage, J. C., Ajenifuja, K. O., Wentzensen, N. A., Adepiti, A. C., Eklund, C., Reilly, M., Hutchinson, M., Wacholder, S., Harford, J., Soliman, A. S., Burk, R. D., and Schiffman, M. (2012). The age-specific prevalence of human papillomavirus and risk of cytologic abnormalities in rural Nigeria: implications for screen-and-treat strategies. *Int J Cancer* **130**, 2111-7.

Garg, A., Suri, V., Nijhawan, R., Aggarwal, N., Aggarwal, R., Guleria, C., and Thakur, M. (2016). Prevalence of Human Papilloma Virus Infection in Young Primiparous Women During Postpartum Period: Study from a Tertiary Care Center in Northern India. *J Clin Diagn Res* **10**, Qc06-qc09.

Ghosh, S., Shetty, R. S., Pattanshetty, S. M., Mallya, S. D., Pandey, D., Kabekkodu, S. P., Kamath, V. G., Prabhu, N., D'Souza, J., and Satyamoorthy, K. (2019). Human papilloma and other DNA virus infections of the cervix: A population based comparative study among tribal and general population in India. *PLoS One* **14**, e0219173.

Ginindza, T. G., Dlamini, X., Almonte, M., Herrero, R., Jolly, P. E., Tsoka-Gwegweni, J. M., Weiderpass, E., Broutet, N., and Sartorius, B. (2017). Prevalence of and associated risk factors for high risk human papillomavirus among sexually active women, Swaziland. *PLoS ONE* **12**.

Giuliano, A. R., Papenfuss, M. R., Denman, C. A., de Zapien, J. G., Abrahamsen, M., and Hunter, J. B. (2005). Human papillomavirus prevalence at the USA-Mexico border among women 40 years of age and older. *Int J STD AIDS* **16**, 247-51.

Gupta, B., Sunnam, L. B., Kumar, A., and Parikipandla, S. (2021). Prevalence of human papillomavirus 16 genotype in Anuppur district, Madhya Pradesh. *Molecular Biology Reports* **48**, 503-511.

Hammouda, D., Clifford, G. M., Pallardy, S., Ayyach, G., Chékiri, A., Boudrich, A., Snijders, P. J., van Kemenade, F. J., Meijer, C. J., Bouhadef, A., Zitouni, Z., Habib, D., Ikezaren, N., and Franceschi, S. (2011). Human papillomavirus infection in a population-based sample of women in Algiers, Algeria. *Int J Cancer* **128**, 2224-9.

Hanisch, R. A., Sow, P. S., Toure, M., Dem, A., Dembele, B., Toure, P., Winer, R. L., Hughes, J. P., Gottlieb, G. S., Feng, Q., Kiviat, N. B., and Hawes, S. E. (2013). Influence of HIV-1 and/or HIV-2 infection and CD4 count on cervical HPV DNA detection in women from Senegal, West Africa. *J Clin Virol* **58**, 696-702.

Hassani, S., Nadji, P. S., Mohseni, A., Rahnamaye Farzami, M., Mirab Samiee, S., Sadr, M., and Nadji, S. A. (2022). Evaluation Frequency of Human Papillomavirus and Its Related Genotypes in Women of the General Population Living in 11 Provinces of Iran. *Can J Infect Dis Med Microbiol* **2022**, 8668557.

Illades-Aguiar, B., Alarcón-Romero Ldel, C., Antonio-Véjar, V., Zamudio-López, N., Sales-Linares, N., Flores-Alfaro, E., Fernández-Tilapa, G., Vences-Velázquez, A., Muñoz-Valle, J. F., and Leyva-Vázquez, M. A. (2010). Prevalence and distribution of human papillomavirus types in cervical cancer, squamous intraepithelial lesions, and with no intraepithelial lesions in women from Southern Mexico. *Gynecol Oncol* **117**, 291-6.

Inal, M. M., Köse, S., Yildirim, Y., Ozdemir, Y., Töz, E., Ertopçu, K., Ozelmas, I., and Tinar, S. (2007). The relationship between human papillomavirus infection and cervical intraepithelial neoplasia in Turkish women. *Int J Gynecol Cancer* **17**, 1266-70.

Kashyap (2013). Value of high-risk human papillomavirus 16 deoxyribonucleic acid testing with cytological entities in peri and postmenopausal women.

Keita, N., Clifford, G. M., Koulibaly, M., Douno, K., Kabba, I., Haba, M., Sylla, B. S., van Kemenade, F. J., Snijders, P. J., Meijer, C. J., and Franceschi, S. (2009). HPV infection in women with and without cervical cancer in Conakry, Guinea. *Br J Cancer* **101**, 202-8.

Khodakarami, N., Clifford, G. M., Yavari, P., Farzaneh, F., Salehpour, S., Broutet, N., Bathija, H., Heideman, D. A. M., Van Kemenade, F. J., Meijer, C. J. L. M., Hosseini, S. J., and Franceschi, S. (2012). Human papillomavirus infection in women with and without cervical cancer in Tehran, Iran. *International Journal of Cancer* **131**, E156-E161.

Krings, A., Dunyo, P., Pesic, A., Tetteh, S., Hansen, B., Gedzah, I., Wormenor, C. M., Amuah, J. E., Behnke, A. L., Höfler, D., Pawlita, M., and Kaufmann, A. M. (2019). Characterization of Human Papillomavirus prevalence and risk factors to guide cervical cancer screening in the North Tongu District, Ghana. *PLoS One* **14**, e0218762.

Laikangbam, P., Sengupta, S., Bhattacharya, P., Duttagupta, C., Dhabali Singh, T., Verma, Y., Roy, S., Das, R., and Mukhopadhyay, S. (2007). A comparative profile of the prevalence and age distribution of human papillomavirus type 16/18 infections among three states of India with focus on northeast India. *International Journal of Gynecological Cancer* **17**, 107-117.

Lazcano-Ponce, E., Herrero, R., Muñoz, N., Cruz, A., Shah, K. V., Alonso, P., Hernández, P., Salmerón, J., and Hernández, M. (2001). Epidemiology of HPV infection among Mexican women with normal cervical cytology. *Int J Cancer* **91**, 412-20.

Levi, J. E., Martins, T. R., Longatto-Filho, A., Cohen, D. D., Cury, L., Fuza, L. M., Villa, L. L., and Eluf-Neto, J. (2019). High-risk HPV testing in primary screening for cervical cancer in the public health system, São Paulo, Brazil. *Cancer Prevention Research* **12**, 539-546.

Leyh-Bannurah, S. R., Prugger, C., de Koning, M. N., Goette, H., and Lellé, R. J. (2014). Cervical human papillomavirus prevalence and genotype distribution among hybrid capture 2 positive women 15 to 64 years of age in the Gurage zone, rural Ethiopia. *Infect Agent Cancer* **9**, 33.

Li, L. K., Dai, M., Clifford, G. M., Yao, W. Q., Arslan, A., Li, N., Shi, J. F., Snijders, P. J., Meijer, C. J., Qiao, Y. L., and Franceschi, S. (2006). Human papillomavirus infection in Shenyang City, People's Republic of China: A population-based study. *Br J Cancer* **95**, 1593-7.

Lippman, S. A., Sucupira, M. C. A., Jones, H. E., Luppi, C. G., Palefsky, J., Van De Wijgert, J. H. H. M., Oliveira, R. L. S., and Diaz, R. S. (2010). Prevalence, distribution and correlates of endocervical human papillomavirus types in Brazilian women. *International Journal of STD and AIDS* **21**, 105-109.

Matos, E., Loria, D., Amestoy, G. M., Herrera, L., Prince, M. A., Moreno, J., Krunfly, C., Van Den Brule, A. J. C., Meijer, C. J. L. M., Muñoz, N., Herrero, R., Converti, N., Fainman, E., Garcia, G., Joannas, S., Kobal, A., Lasco, M. S., Ledo, P., Mas, A., Oppel, E., Ragone, M. A., Rios, H., Ripoll, M. A., Rivas, F., Rivas, J., Rivero, G., Rodriquez, G., Scattone, C., Tolomei, R., Rodriguez, A. M., Taborda, M. F., Lorenzo, Y., Barrios, L., Pellandino, M., Quiroz, C., and Vilensky, M. (2003). Prevalence of human papillomavirus infection among women in Concordia, Argentina: A population-based study. *Sexually Transmitted Diseases* **30**, 593-599.

Mbulawa, Z. Z. A., Marais, D. J., Johnson, L. F., Boulle, A., Coetzee, D., and Williamson, A. L. (2010). Influence of human immunodeficiency virus and CD4 count on the prevalence of human papillomavirus in heterosexual couples. *Journal of General Virology* **91**, 3023-3031.

Miranda, P. M., Pitol, B. C., Moran, M. S., Silva, N. N., Felix, P. M., Lima-Filho, J. L., Carneiro, C. M., Silva, I. D., Carvalho, R. F., Lima, A. A., Beçak, W., and Stocco, R. C. (2012). Human papillomavirus infection in Brazilian women with normal cervical cytology. *Genet Mol Res* **11**, 1752-61.

Molano, M., Posso, H., Weiderpass, E., Van den Brule, A. J. C., Ronderos, M., Franceschi, S., Meijer, C. J. L. M., Arslan, A., and Munoz, N. (2002). Prevalence and determinants of HPV infection among Colombian women with normal cytology. *British Journal of Cancer* **87**, 324-333.

Nahar, Q., Sultana, F., Alam, A., Islam, J. Y., Rahman, M., Khatun, F., Alam, N., Dasgupta, S. K., Marions, L., Ashrafunnessa, Kamal, M., Cravioto, A., and Reichenbach, L. (2014). Genital human papillomavirus infection among women in Bangladesh: Findings from a population-based survey. *PLoS ONE* **9**.

Ngabo, F., Franceschi, S., Baussano, I., Umulisa, M. C., Snijders, P. J. F., Uyterlinde, A. M., Lazzarato, F., Tenet, V., Gatera, M., Binagwaho, A., and Clifford, G. M. (2016). Human papillomavirus infection in Rwanda at the moment of implementation of a national HPV vaccination programme. *BMC Infectious Diseases* **16**.

Oliveira, L. H., Ferreira, M. D., Augusto, E. F., Melgaço, F. G., Santos, L. S., Cavalcanti, S. M., and Rosa, M. L. (2010). Human papillomavirus genotypes in asymptomatic young women from public schools in Rio de Janeiro, Brazil. *Rev Soc Bras Med Trop* **43**, 4-8.

Phongsavan, K., Gustavsson, I., Marions, L., Phengsavanh, A., Wahlström, R., and Gyllensten, U. (2012). Detection of human papillomavirus among women in Laos: feasibility of using filter paper card and prevalence of high-risk types. *Int J Gynecol Cancer* **22**, 1398-406.

Ramatlho, P., Grover, S., Mathoma, A., Tawe, L., Matlhagela, K., Ngoni, K., Molebatsi, K., Chilisa, B., Zetola, N. M., Robertson, E. S., Paganotti, G. M., and Ramogola-Masire, D. (2022). Human papillomavirus prevalence among unvaccinated young female college students in Botswana: A cross-sectional study. *S Afr Med J* **112**, 335-340.

Raza, S. A., Franceschi, S., Pallardy, S., Malik, F. R., Avan, B. I., Zafar, A., Ali, S. H., Pervez, S., Serajuddaula, S., Snijders, P. J., van Kemenade, F. J., Meijer, C. J., Shershah, S., and Clifford, G. M. (2010). Human papillomavirus infection in women with and without cervical cancer in Karachi, Pakistan. *Br J Cancer* **102**, 1657-60.

Richter, K., Becker, P., Horton, A., and Dreyer, G. (2013). Age-specific prevalence of cervical human papillomavirus infection and cytological abnormalities in women in Gauteng Province, South Africa. *S Afr Med J* **103**, 313-7.

Safaei, A., Khanlari, M., Momtahen, M., Monabati, A., Robati, M., Amooei, S., Valibeigi, B., and Azarpira, N. (2010). Prevalence of high-risk human papillomavirus types 16 and 18 in healthy women with cytologically negative pap smear in Iran. *Indian Journal of Pathology and Microbiology* **53**, 681-685.

Safaeian, M., Herrero, R., Hildesheim, A., Quint, W., Freer, E., Van Doorn, L. J., Porras, C., Silva, S., González, P., Bratti, M. C., Rodriguez, A. C., and Castle, P. (2007a). Comparison of the SPF10-LiPA system to the Hybrid Capture 2 Assay for detection of carcinogenic human papillomavirus genotypes among 5,683 young women in Guanacaste, Costa Rica. *J Clin Microbiol* **45**, 1447-54.

Safaeian, M., Kiddugavu, M., Gravitt, P. E., Ssekasanvu, J., Murokora, D., Sklar, M., Serwadda, D., Wawer, M. J., Shah, K. V., and Gray, R. (2007b). Comparability of self-collected vaginal swabs and physician-collected cervical swabs for detection of human papillomavirus infections in Rakai, Uganda. *Sex Transm Dis* **34**, 429-36.

Shakya, S., Syversen, U., Åsvold, B. O., Bofin, A. M., Aune, G., Nordbø, S. A., Vaidya, K. M., Karmacharya, B. M., Afset, J. E., and Tingulstad, S. (2017). Prevalence of human papillomavirus infection among women in rural Nepal. *Acta Obstetricia et Gynecologica Scandinavica* **96**, 29-38.

Sherpa, A. T. L., Clifford, G. M., Vaccarella, S., Shrestha, S., Nygard, M., Karki, B. S., Snijders, P. J. F., Meijer, C. J. L. M., and Franceschi, S. (2010). Human papillomavirus infection in women with and without cervical cancer in Nepal. *Cancer Causes and Control* **21**, 323-330.

Sowjanaya, A. P., Jain, M., Poli, U. R., Padma, S., Das, M., Shah, K. V., Rao, B. N., Devi, R. R., Gravitt, P. E., and Ramakrishna, G. (2005). Prevalence and distribution of high-risk human papilloma virus (HPV) types in invasive squamous cell carcinoma of the cervix and in normal women in Andhra Pradesh, India. *BMC Infectious Diseases* **5**.

Sriamporn, S., Snijders, P. J., Pientong, C., Pisani, P., Ekalaksananan, T., Meijer, C. J., and Parkin, D. M. (2006). Human papillomavirus and cervical cancer from a prospective study in Khon Kaen, Northeast Thailand. *Int J Gynecol Cancer* **16**, 266-9.

Subramanian, M. J., Rajaraman, S., and Vijayakumar, V. (2021). A Community-Based Study on Prevalence, Genotype Distribution and Persistence of High-Risk Human Papilloma Virus Infection of Uterine Cervix in Rural South India. *Indian Journal of Gynecologic Oncology* **19**.

Sukvirach, S., Smith, J. S., Tunsakul, S., Muñoz, N., Kesararat, V., Opasatian, O., Chichareon, S., Kaenploy, V., Ashley, R., Meijer, C. J., Snijders, P. J., Coursaget, P., Franceschi, S., and Herrero, R. (2003). Population-based human papillomavirus prevalence in Lampang and Songkla, Thailand. *J Infect Dis* **187**, 1246-56.

Tábora, N., Zelaya, A., Bakkers, J., Melchers, W. J. G., and Ferrera, A. (2005). Chlamydia trachomatis and genital human papillomavirus infections in female university students in Honduras. *American Journal of Tropical Medicine and Hygiene* **73**, 50-53.

Tamegão-Lopes, B. P., Sousa-Júnior, E. C., Passetti, F., Ferreira, C. G., De Mello, W. A., and Duarte Silvestre, R. V. (2014). Prevalence of human papillomavirus infection and phylogenetic analysis of HPV-16 E6 variants among infected women from Northern Brazil. *Infectious Agents and Cancer* **9**.

Thomas, J. O., Herrero, R., Omigbodun, A. A., Ojemakinde, K., Ajayi, I. O., Fawole, A., Oladepo, O., Smith, J. S., Arslan, A., Muñoz, N., Snijders, P. J., Meijer, C. J., and Franceschi, S. (2004). Prevalence of papillomavirus infection in women in Ibadan, Nigeria: a population-based study. *Br J Cancer* **90**, 638-45.

Tshomo, U., Franceschi, S., Dorji, D., Baussano, I., Tenet, V., Snijders, P. J., Meijer, C. J., Bleeker, M. C., Gheit, T., Tommasino, M., and Clifford, G. M. (2014). Human papillomavirus infection in Bhutan at the moment of implementation of a national HPV vaccination programme. *BMC Infect Dis* **14**, 408.

Vidal, A., Murphy, S. K., Hernandez, B., Oneko, O., Overcash, F., and Hoyo, C. (2011). Distribution of HPV genotypes in cervical intraepithelial lesions and cervical cancer in Tanzanian women. *Cancer Prevention Research* **4**.

Vora, K. S., Saiyed, S., Joshi, R., and Natesan, S. (2023). Prevalence of high-risk HPV among marginalized urban women in India and its implications on vaccination: A cross sectional study. *International Journal of Gynecology and Obstetrics* **162**, 176-182.

Vu (2012). "High-risk and multiple human papillomavirus infections among married women in Can Tho, Viet Nam".

Watson-Jones, D., Baisley, K., Brown, J., Kavishe, B., Andreasen, A., Changalucha, J., Mayaud, P., Kapiga, S., Gumodoka, B., Hayes, R. J., and de Sanjosé, S. (2013). High prevalence and incidence of human papillomavirus in a cohort of healthy young African female subjects. *Sex Transm Infect* **89**, 358-65.

Watt, A., Garwood, D., Jackson, M., Younger, N., Ragin, C., Smikle, M., Fletcher, H., and McFarlane-Anderson, N. (2009). High-risk and multiple human papillomavirus (HPV) infections in cancer-free Jamaican women. *Infect Agent Cancer* **4 Suppl 1**, S11.

Wongworapat, K., Keawvichit, R., Sirirojn, B., Dokuta, S., Ruangyuttikarn, C., Sriplienchan, S., Sontirat, A., Kla, K. T., Gravitt, P. E., and Celentano, D. D. (2008). Detection of human papillomavirus from self-collected vaginal samples of women in Chiang Mai, Thailand. *Sex Transm Dis* **35**, 172-3.

Wu, R. F., Dai, M., Qiao, Y. L., Clifford, G. M., Liu, Z. H., Arslan, A., Li, N., Shi, J. F., Snijders, P. J., Meijer, C. J., and Franceschi, S. (2007). Human papillomavirus infection in women in Shenzhen City, People's Republic of China, a population typical of recent Chinese urbanisation. *Int J Cancer* **121**, 1306-11.

Xi, L. F., Touré, P., Critchlow, C. W., Hawes, S. E., Dembele, B., Sow, P. S., and Kiviat, N. B. (2003). Prevalence of specific types of human papillomavirus and cervical squamous intraepithelial lesions in consecutive, previously unscreened, West-African women over 35 years of age. *Int J Cancer* **103**, 803-9.

### 4 Supplementary figures

**eFigure 1. Contributions of the DHS sexual behavior indicators to the six main dimensions identified by the Principal Component Analysis.**
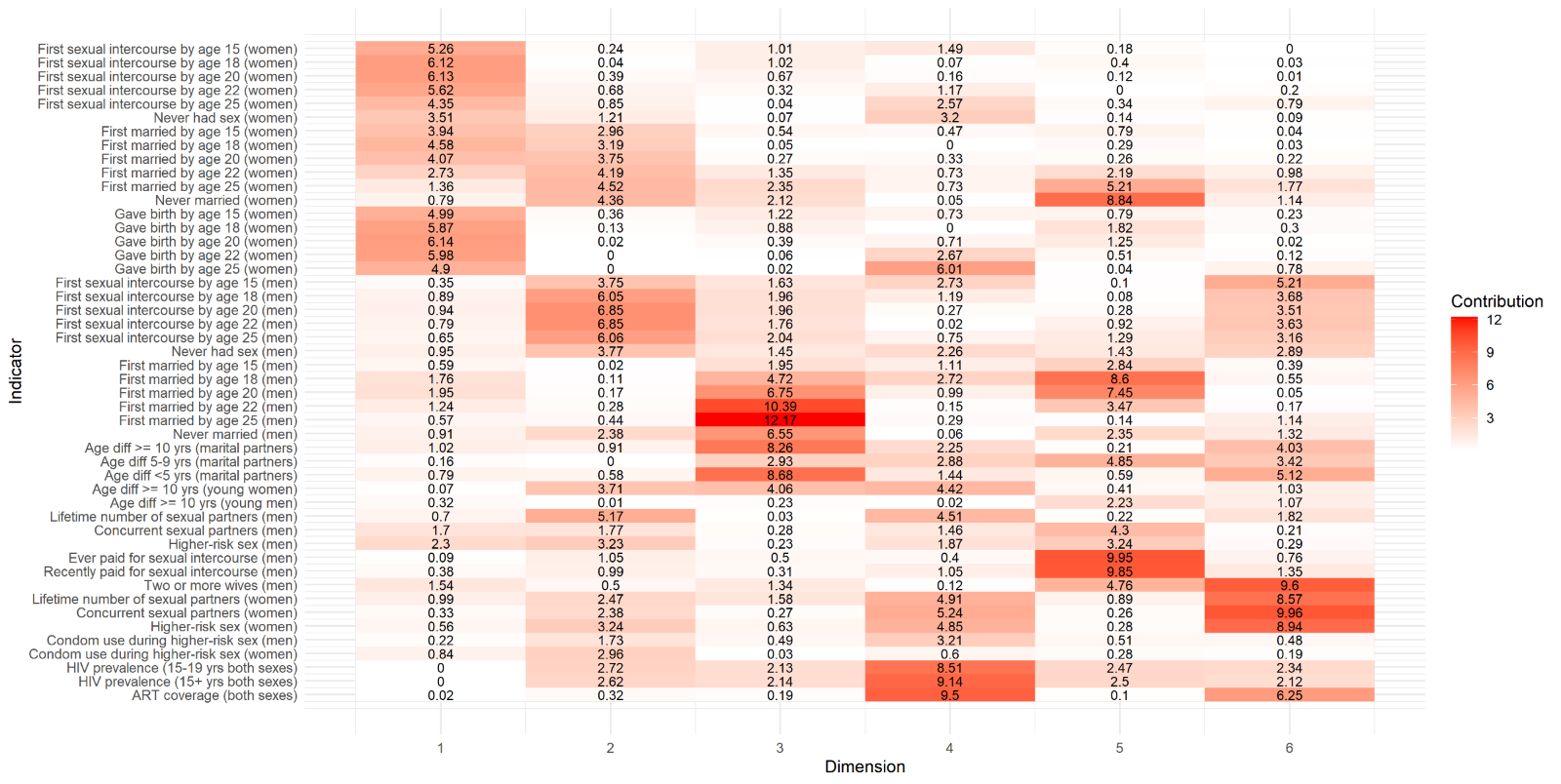

**eFigure 2. Correlation between the 45 original DHS sexual behavior indicators included in the analysis.**

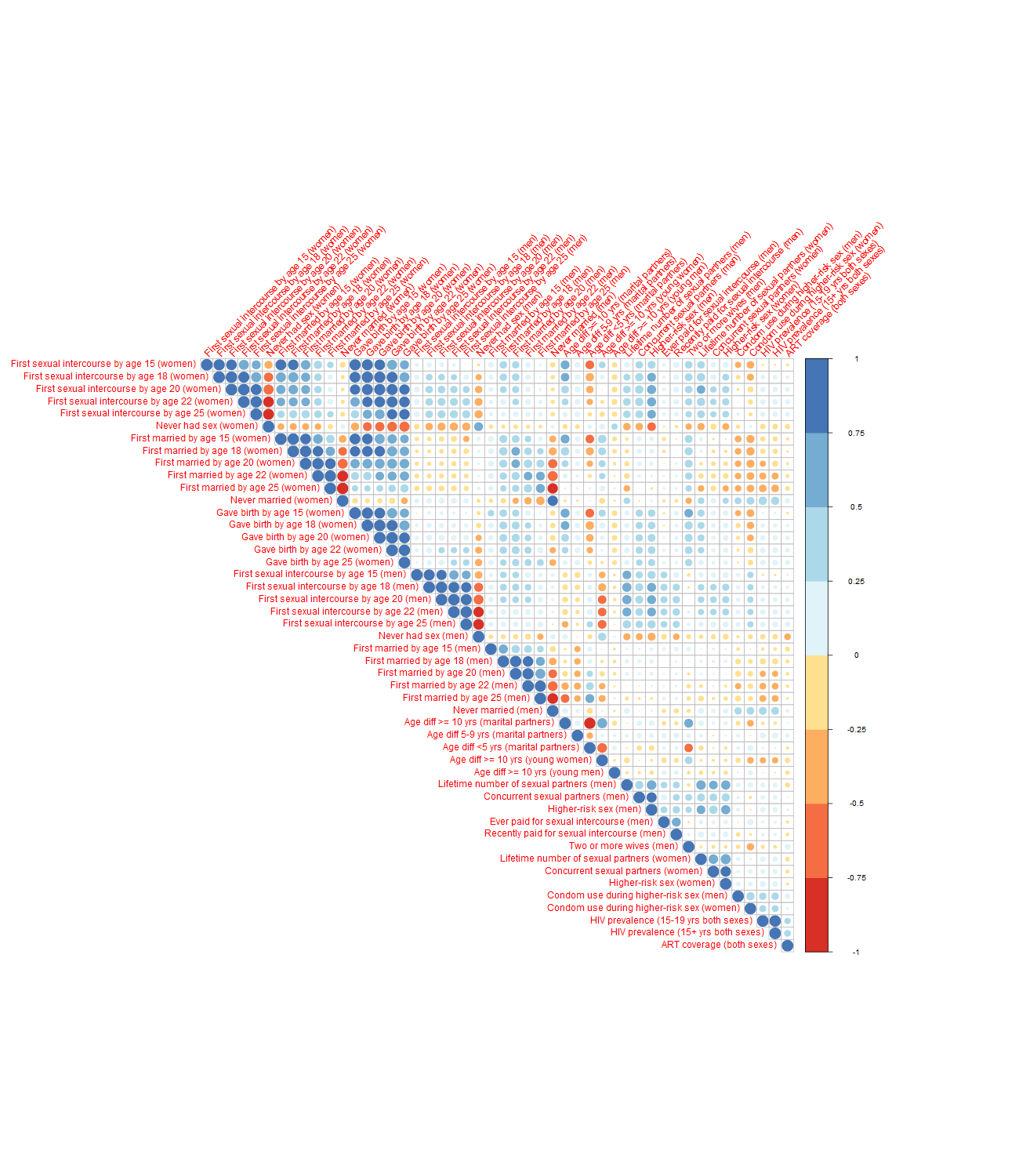

**eFigure 3. Screeplot of Principal Component Analysis showing the percentage of the total variance explained by dimension.**

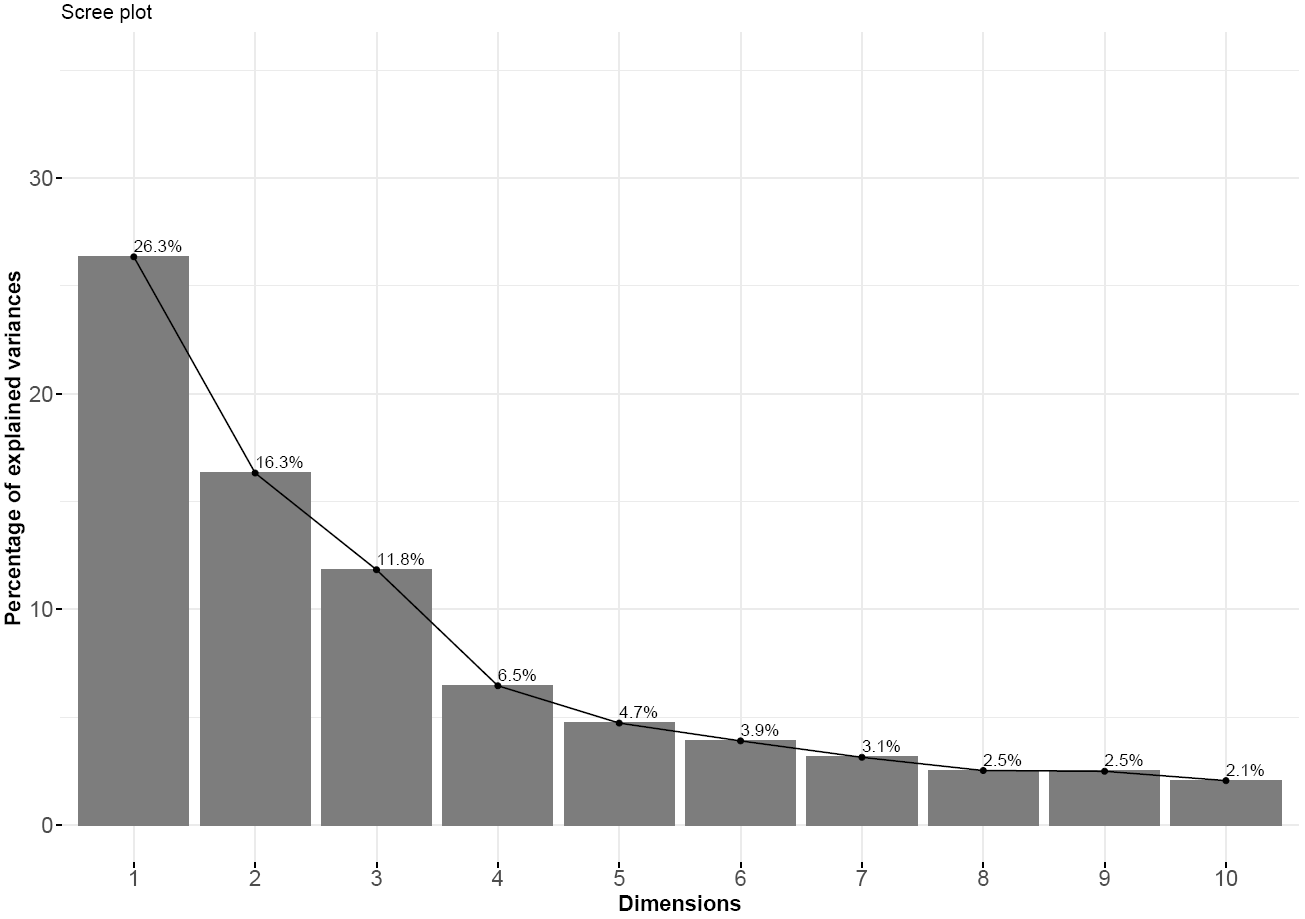

**eFigure 4. Comparison of 4- and 5-clustering in the six dimensions.**

|  | **Dimensions 1 and 2** | **Dimensions 3 and 4** | **Dimensions 5 and 6** |
| --- | --- | --- | --- |
| **4-clustering** | 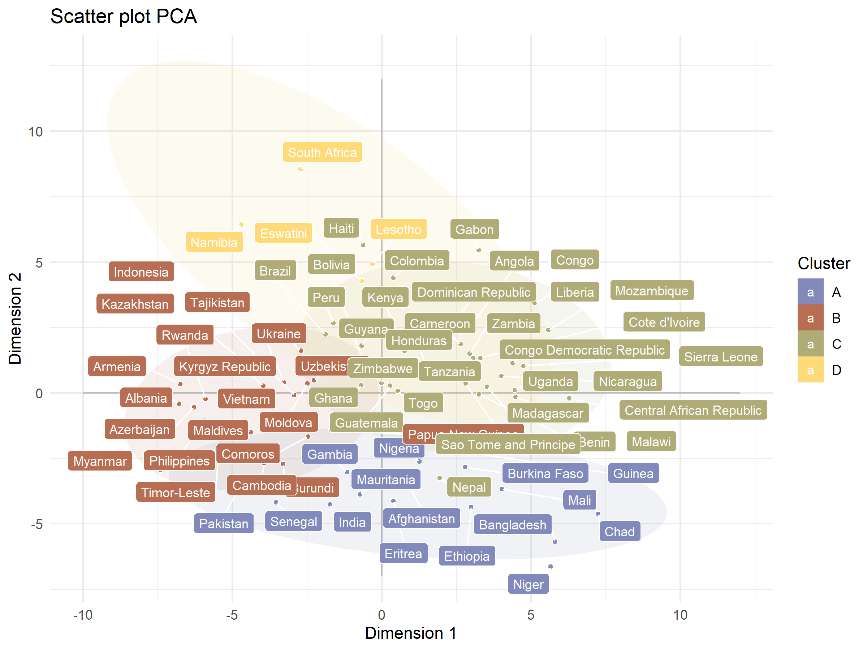 | 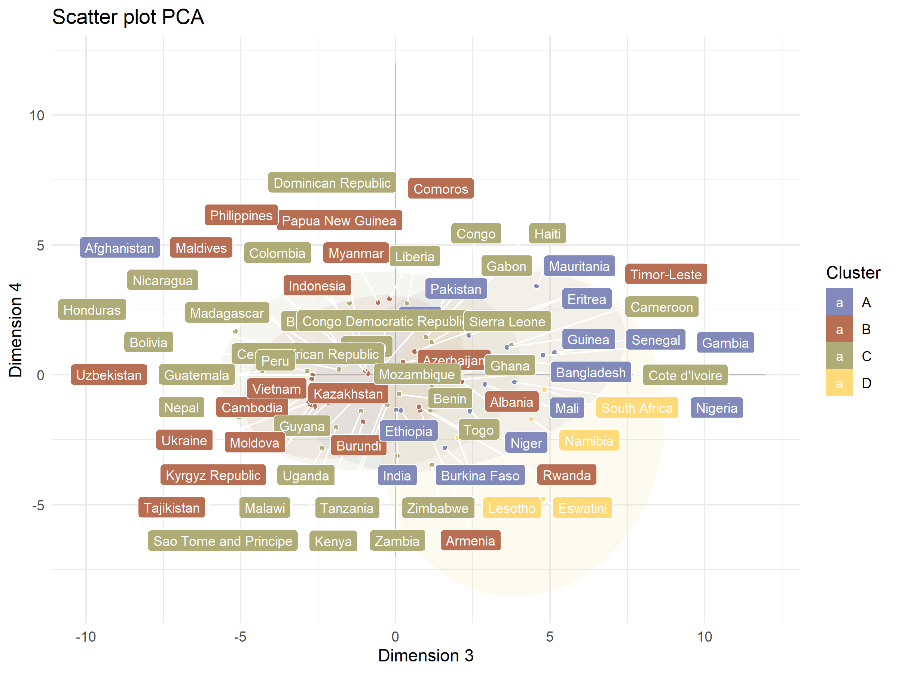 | 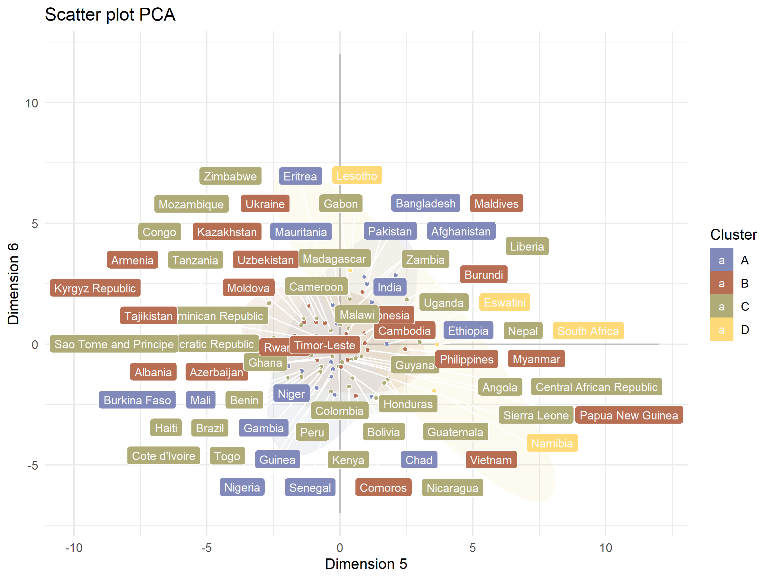 |
| **5-clustering** | 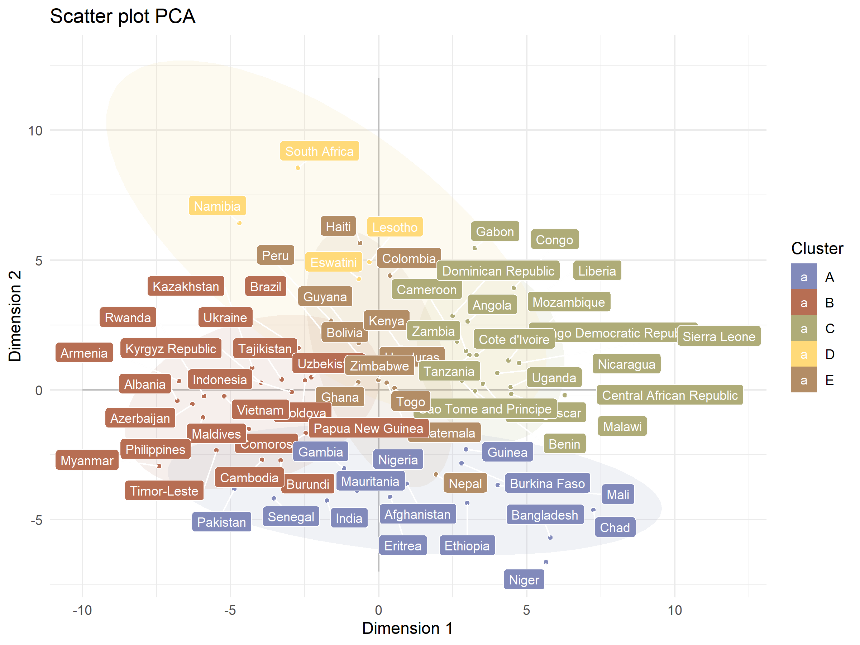 | 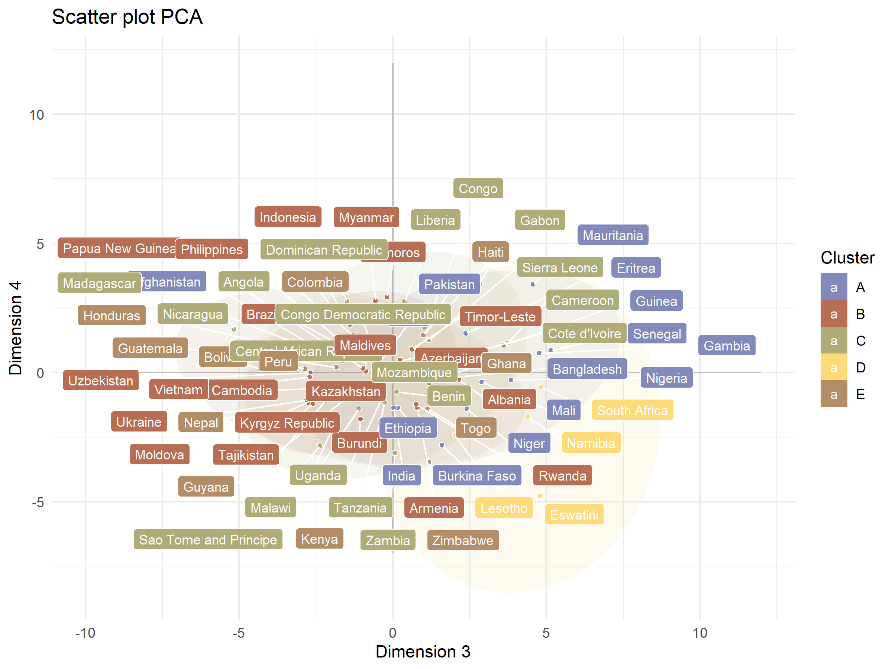 | 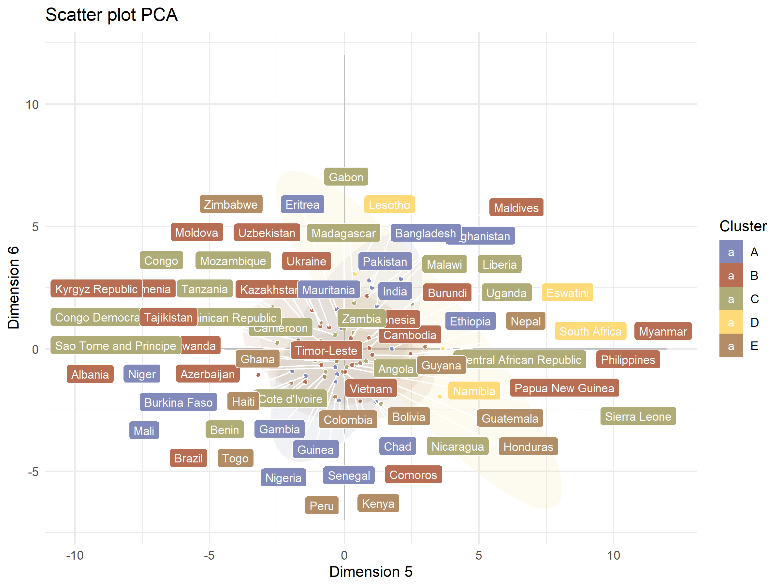 |

**eFigure 5. Comparison of 4- and 5-clustering in the six dimensions in the world map.**

| **4-clustering** | 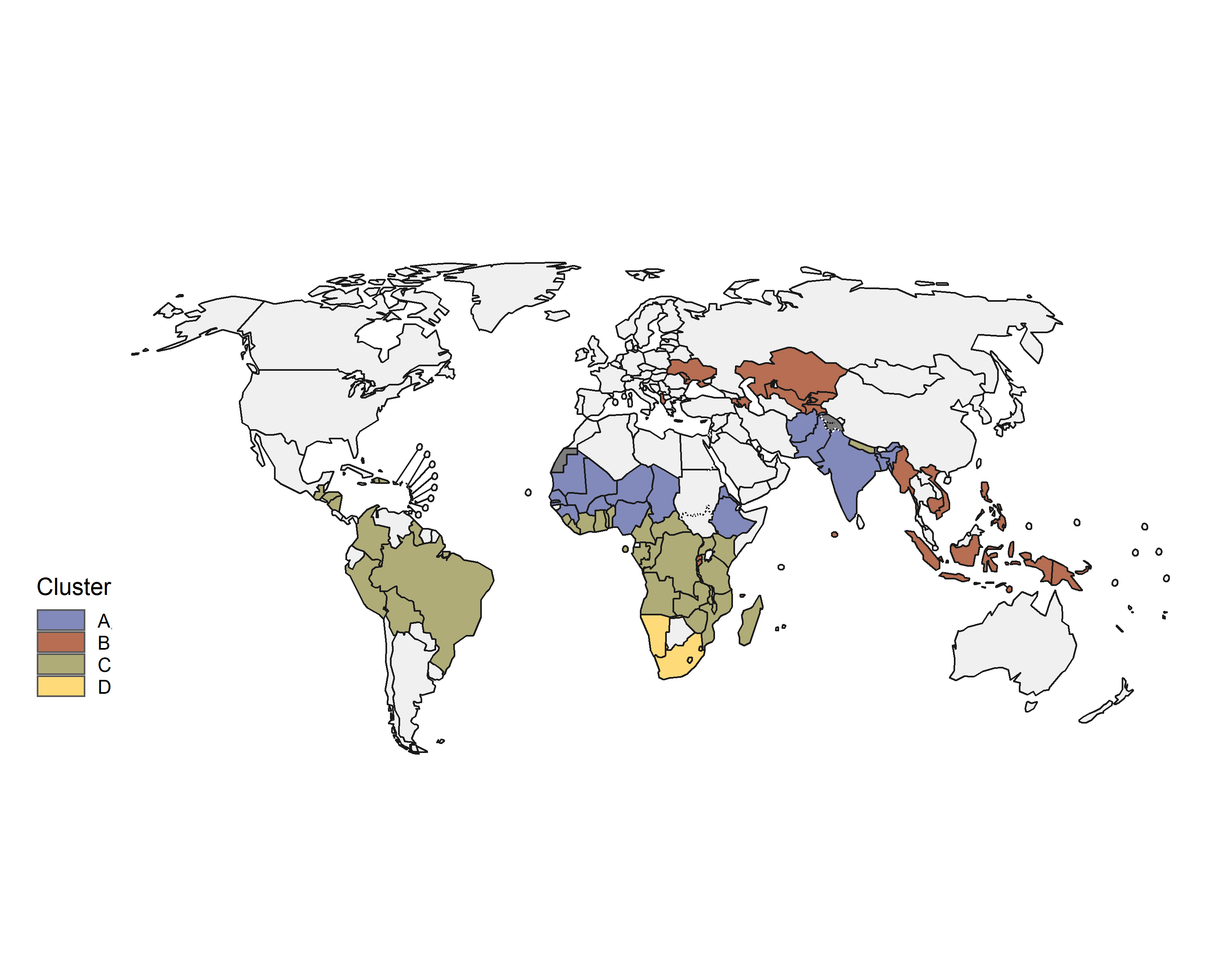 |
| --- | --- |
| **5-clustering** | 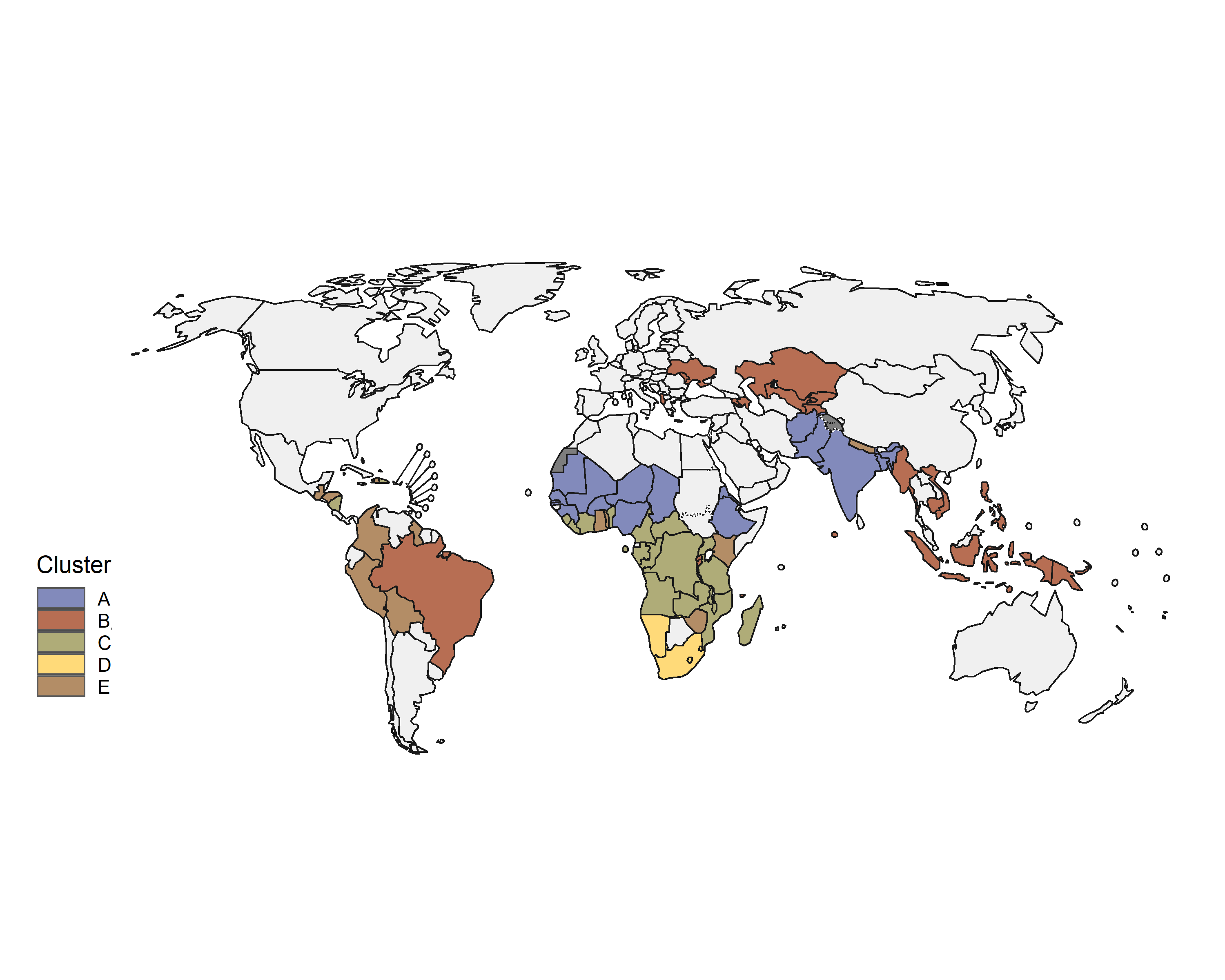 |

**eFigure 6. Results of the 4-clustering in the six dimensions**
The 4 main colors (purple, red, green, and yellow) represent the 4 main clusters, while different shades of the same color represent the region-specific subclusters. The coordinates of each country on the plane are the median coordinates of its 500 imputed representations. The ellipses represent the multivariate student distribution for each cluster. The table below shows the values of some key sexual behavior indicators (ordered by PCA dimension) and the cervical cancer incidence. W=Woman; M=Man.

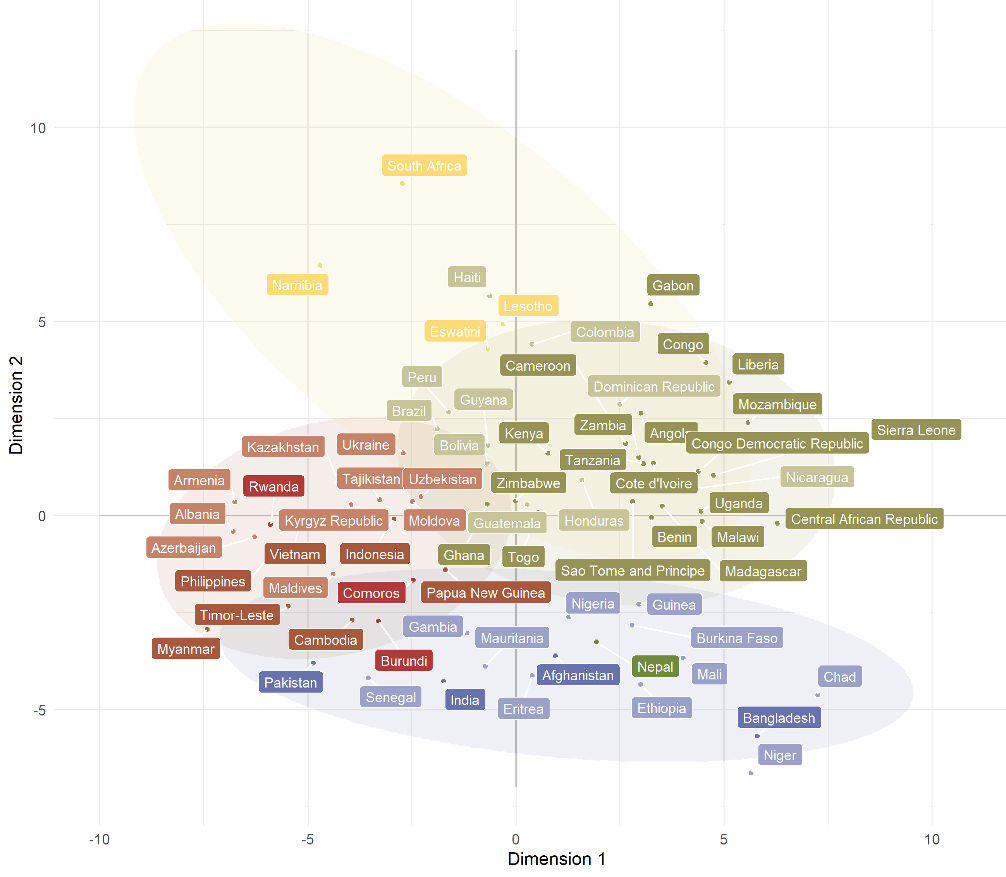

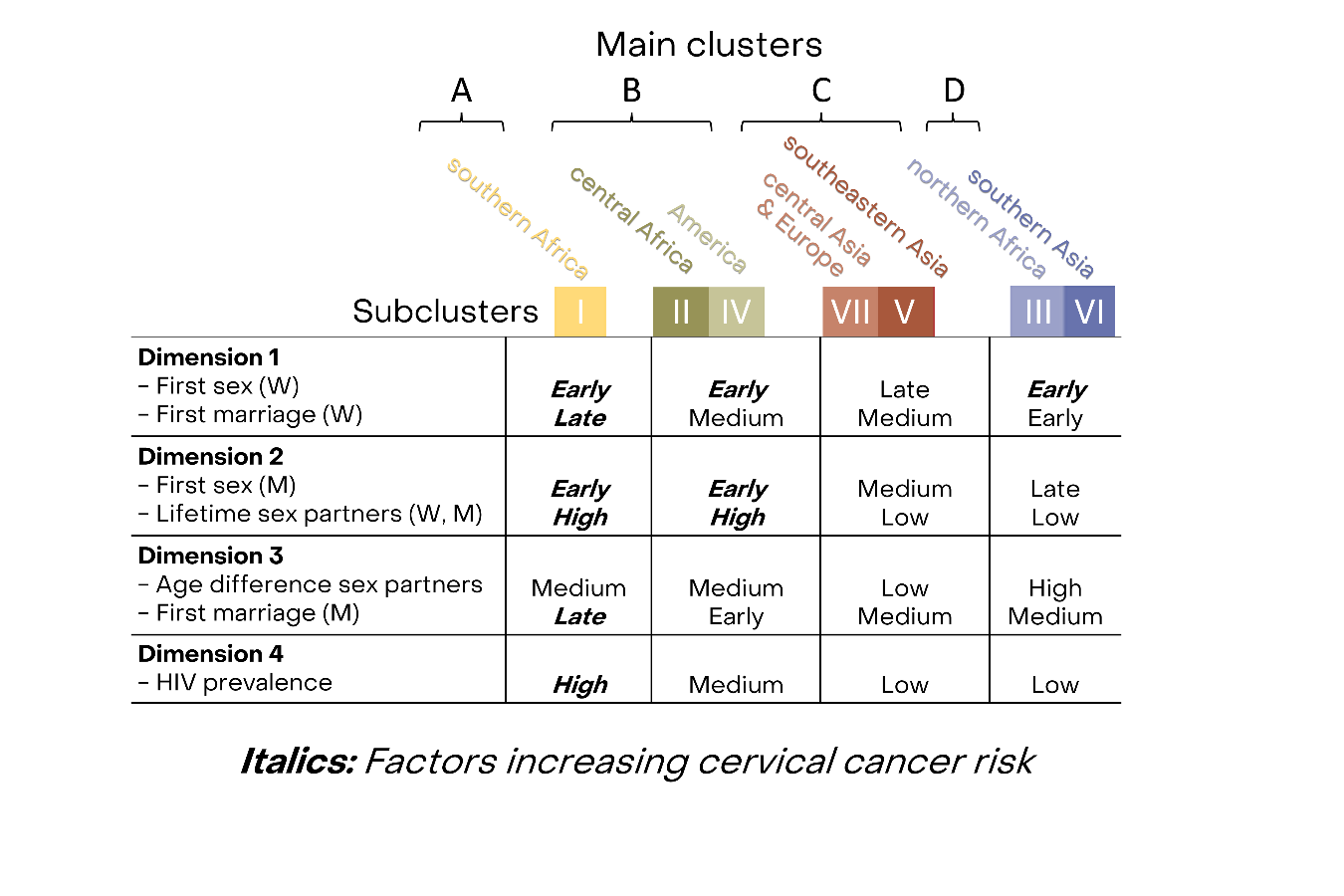

**eFigure 7. Average cervical cancer incidence by cluster.**
ASIR = Aged-standardized incidence rate of cervical cancer in cases per 100,000 women per year.

Average of cervical cancer incidence by cluster

Incidence in cases per 100,000 women per year

|  | Mean | Median |
| --- | --- | --- |
| I | 55.7525 | 46.97 |
| II | 39.0182 | 33.01 |
| III | 28.8725 | 27.235 |
| IV | 21.03 | 19.5 |
| V | 17.0267 | 15.51 |
| VI | 11.762 | 11.28 |
| VII | 11.5822 | 14.14 |

**eFigure 8. Mean number of new partners per year by cluster and by sex based on DHS data**.
The black lines represent the average for all classes of sexual activity (CSA). CSA h=high, l=low, m=medium (the assumed proportion of people belonging to each CSA in the population is assumed to be 0.05, 0.15, and 0.8 for high, medium, and low CSA, respectively. These sexual behaviour parameters were used in HPV transmission model RHEA, and were constructed based on DHS data. M=men, W=women.

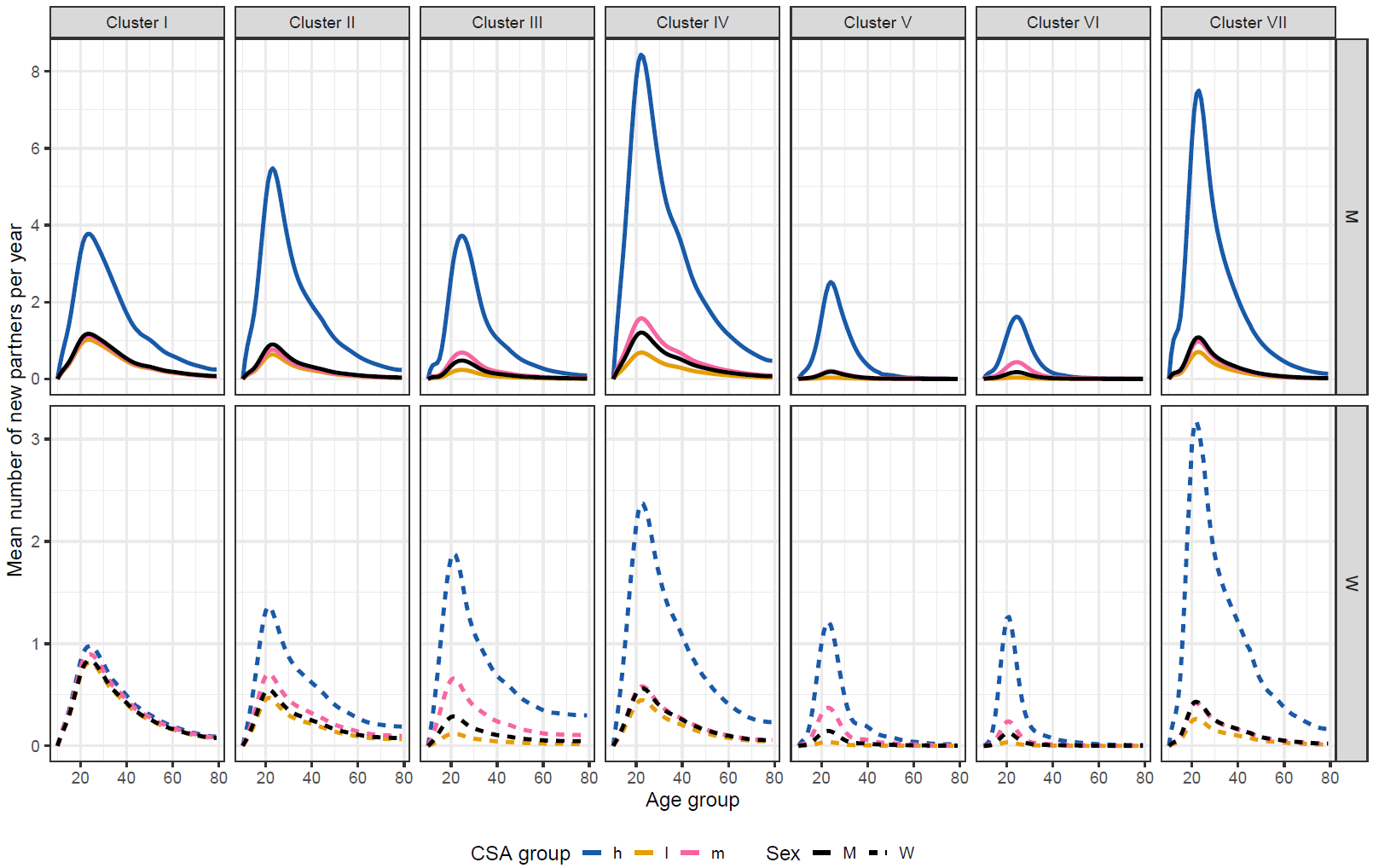

**eFigure 9. Mixing patterns in women by age and age of the male partner based on DHS data.**
Patterns are given by distribution of the age of the male partner given the age of women. Patterns are shown by cluster. These sexual behaviour parameters were used in HPV transmission model RHEA and were constructed based on DHS data.

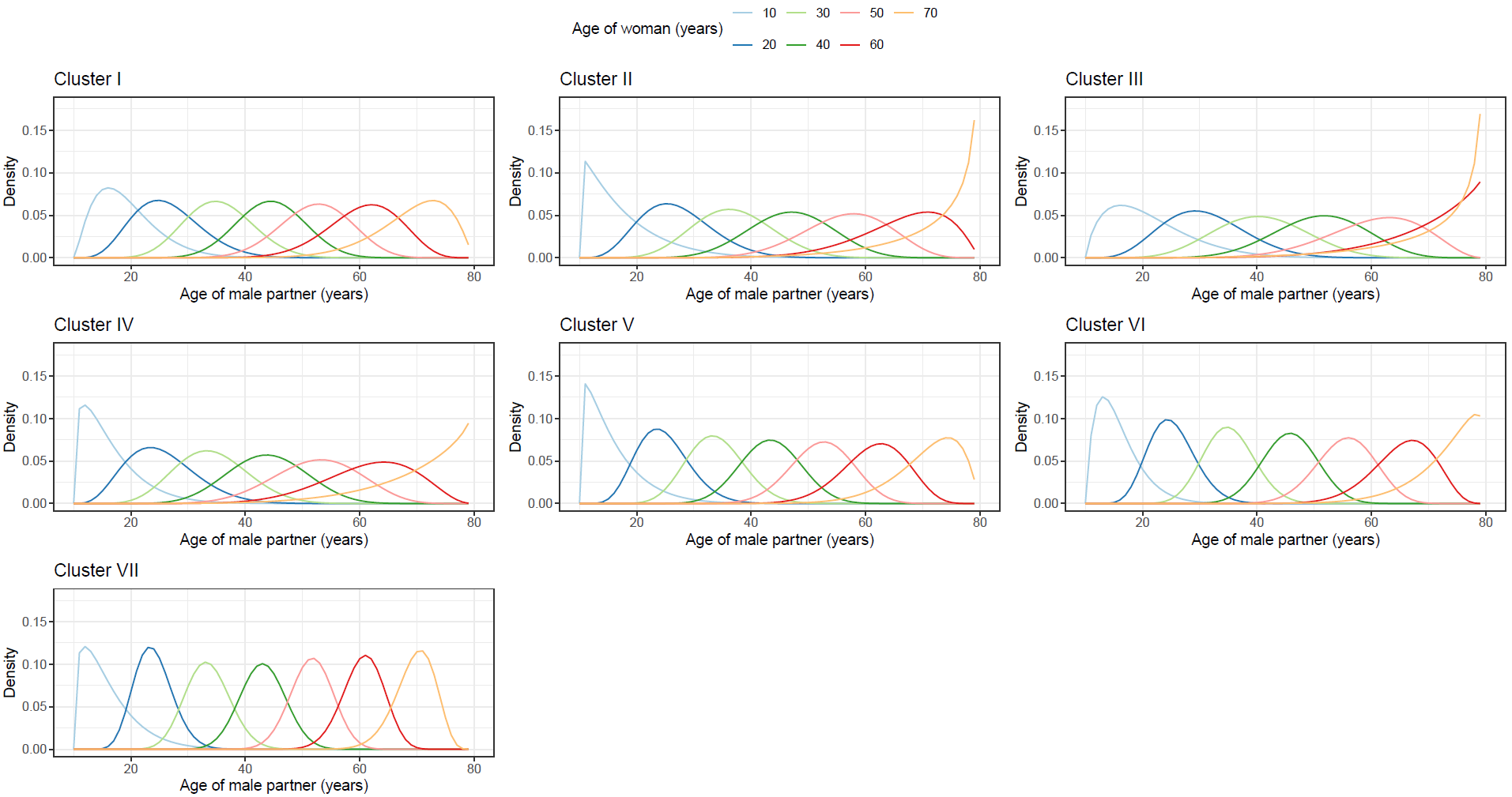

**eFigure 10. Calibrated sexual parameters and infection transmission probabilities of the HPV transmission model RHEA.**
The first four panels are the four type-specific transmission probabilities, here called beta, by groups of high-risk HPV type: 16, 18, 31/33/45/52/58 (Nona), and other high-risk types. The second four panels are the two assortativeness adjustment parameters, by csa and sex (m=men, w=women), and by age and sex. Values close to 1 correspond to more proportionate mixing whereas values close to 0 correspond to more assortative mixing within the same group. The last two panels are the two adjustment parameters to account for under/over-reporting of sexual contact rate in the DHS (one adjustment parameter for ages 10-19, and one for ages 20-79). High values correspond to more underreporting.

**Cluster I**

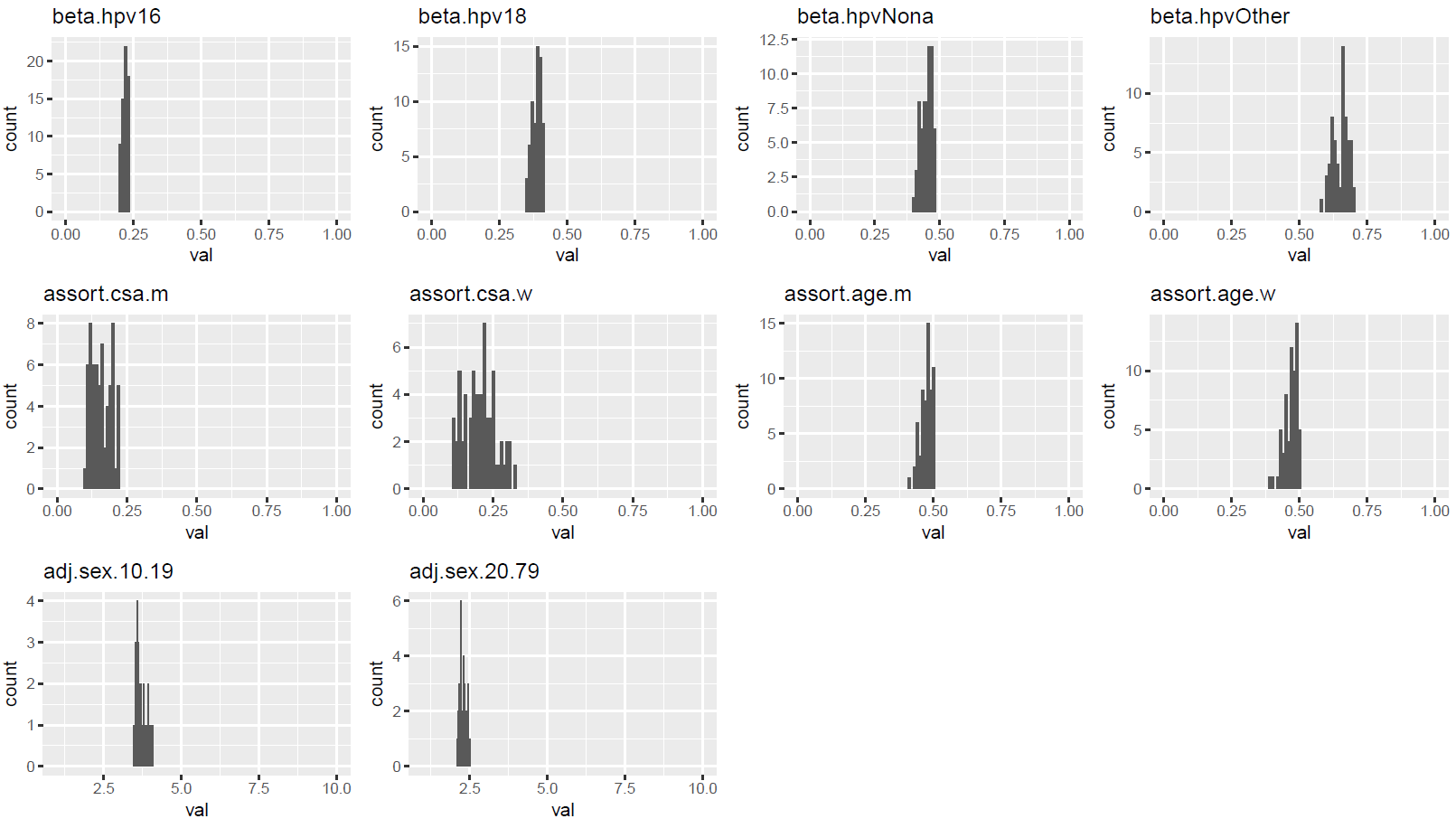

**Cluster II**

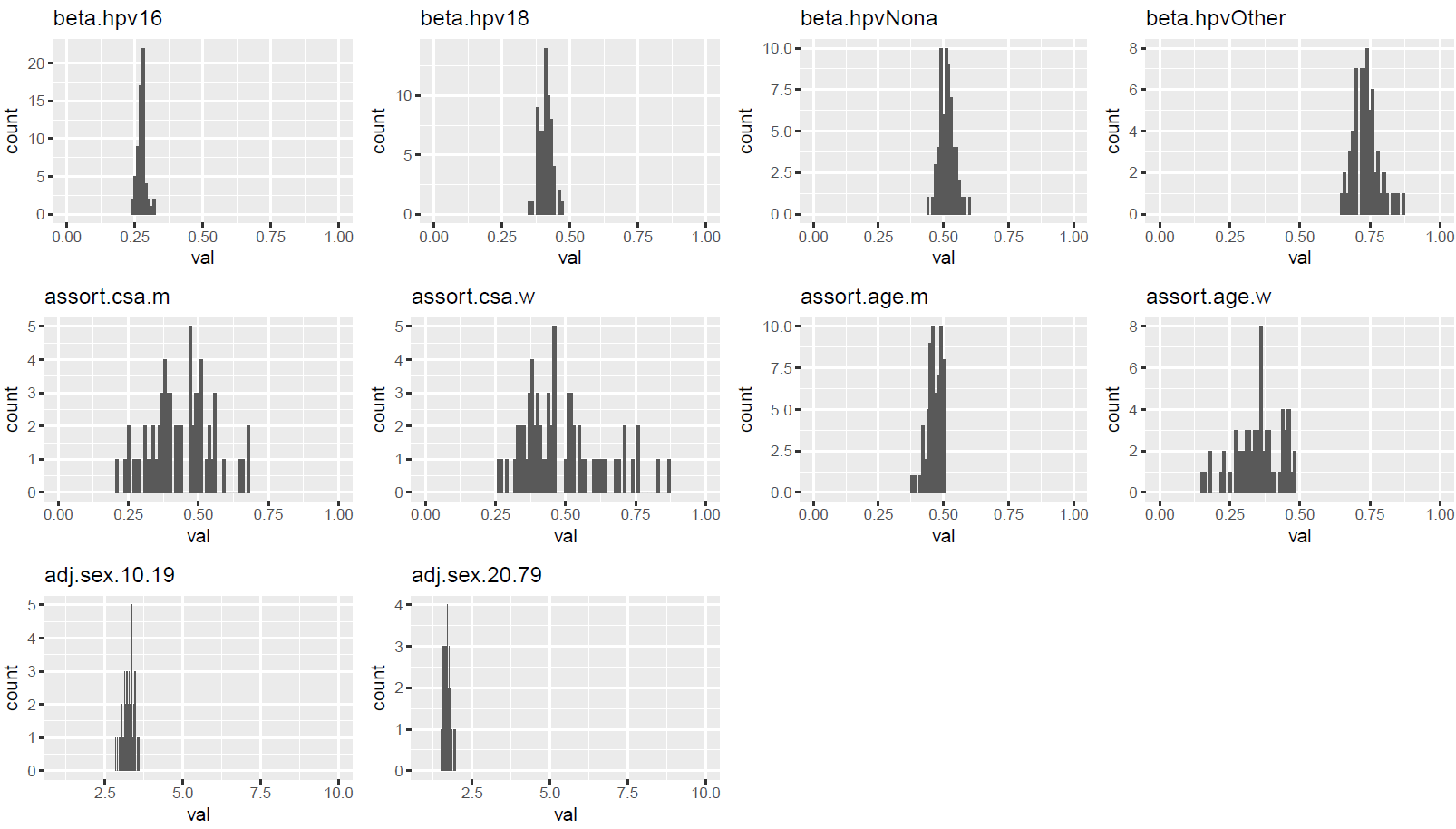

**Cluster III**

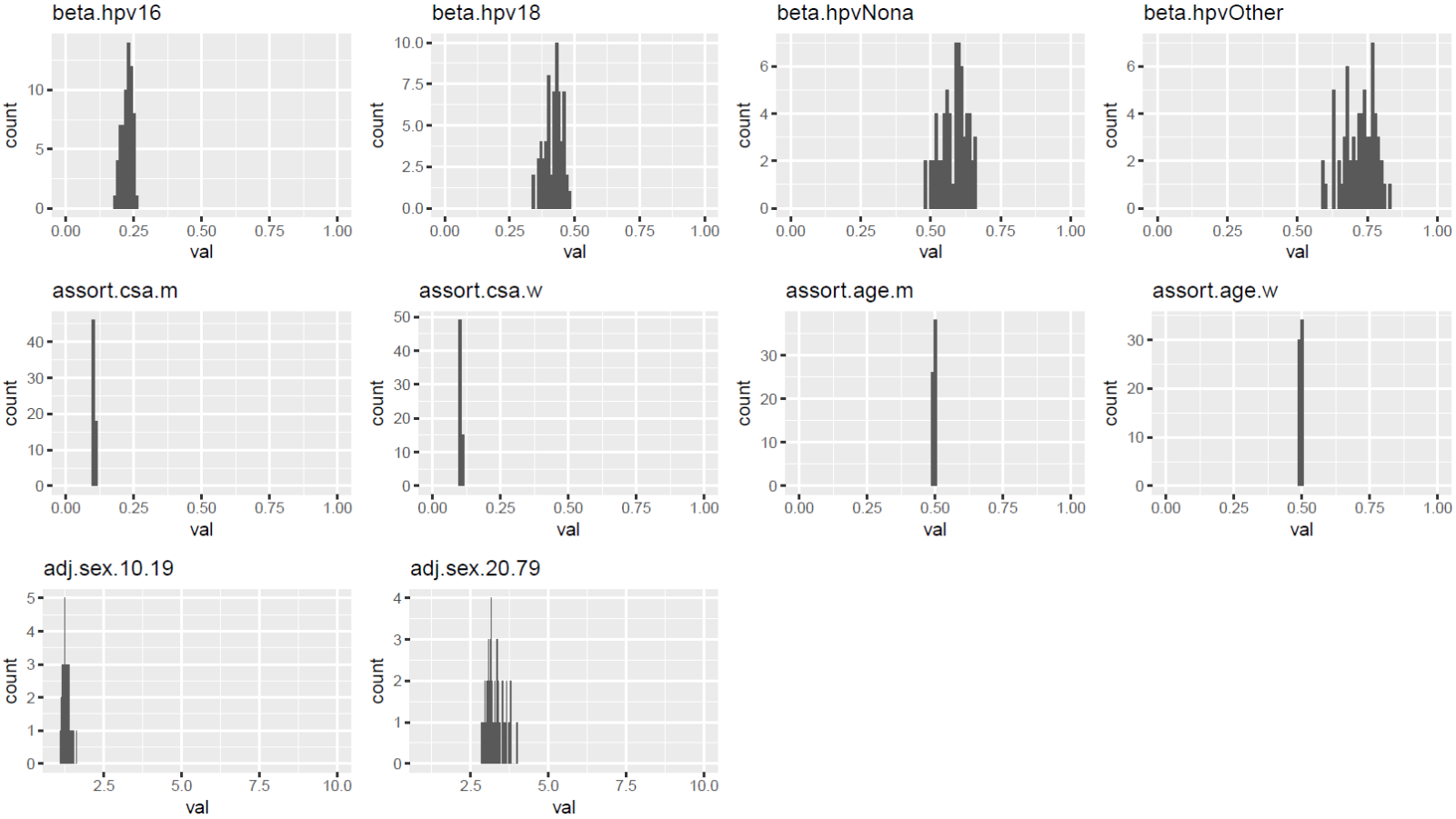

**Cluster IV**

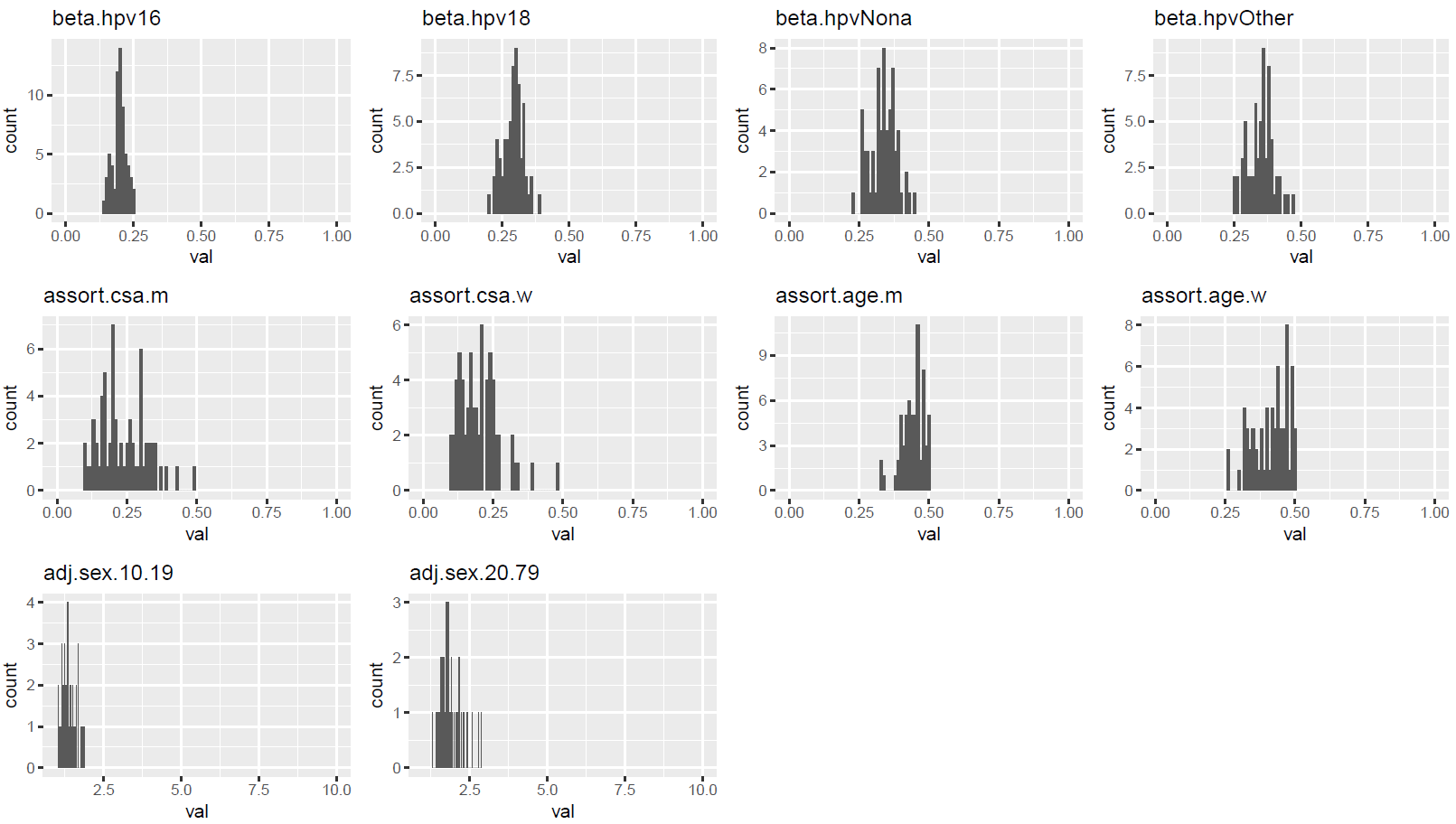

**Cluster V**

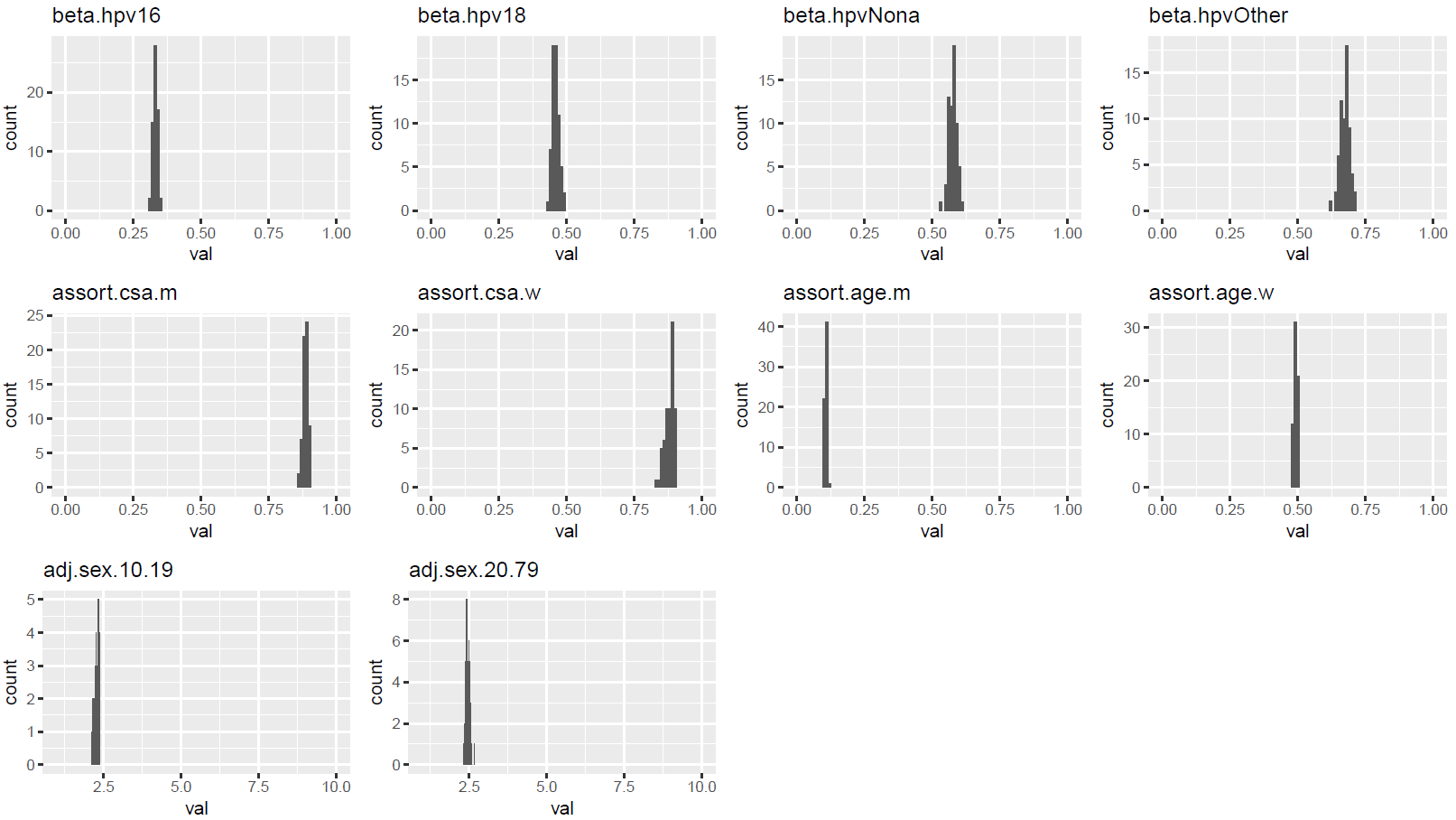

**Cluster VI**

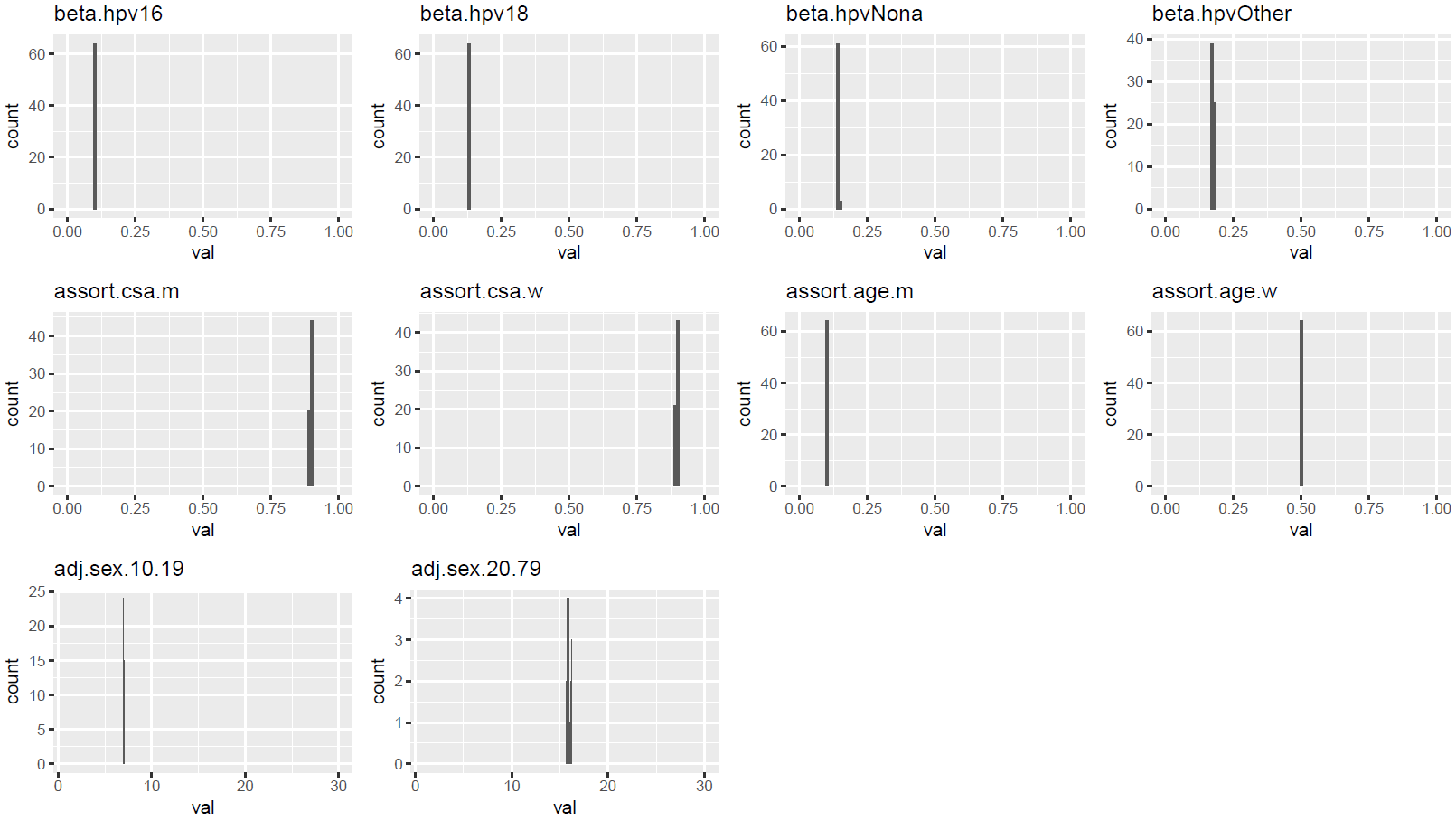

**Cluster VII**

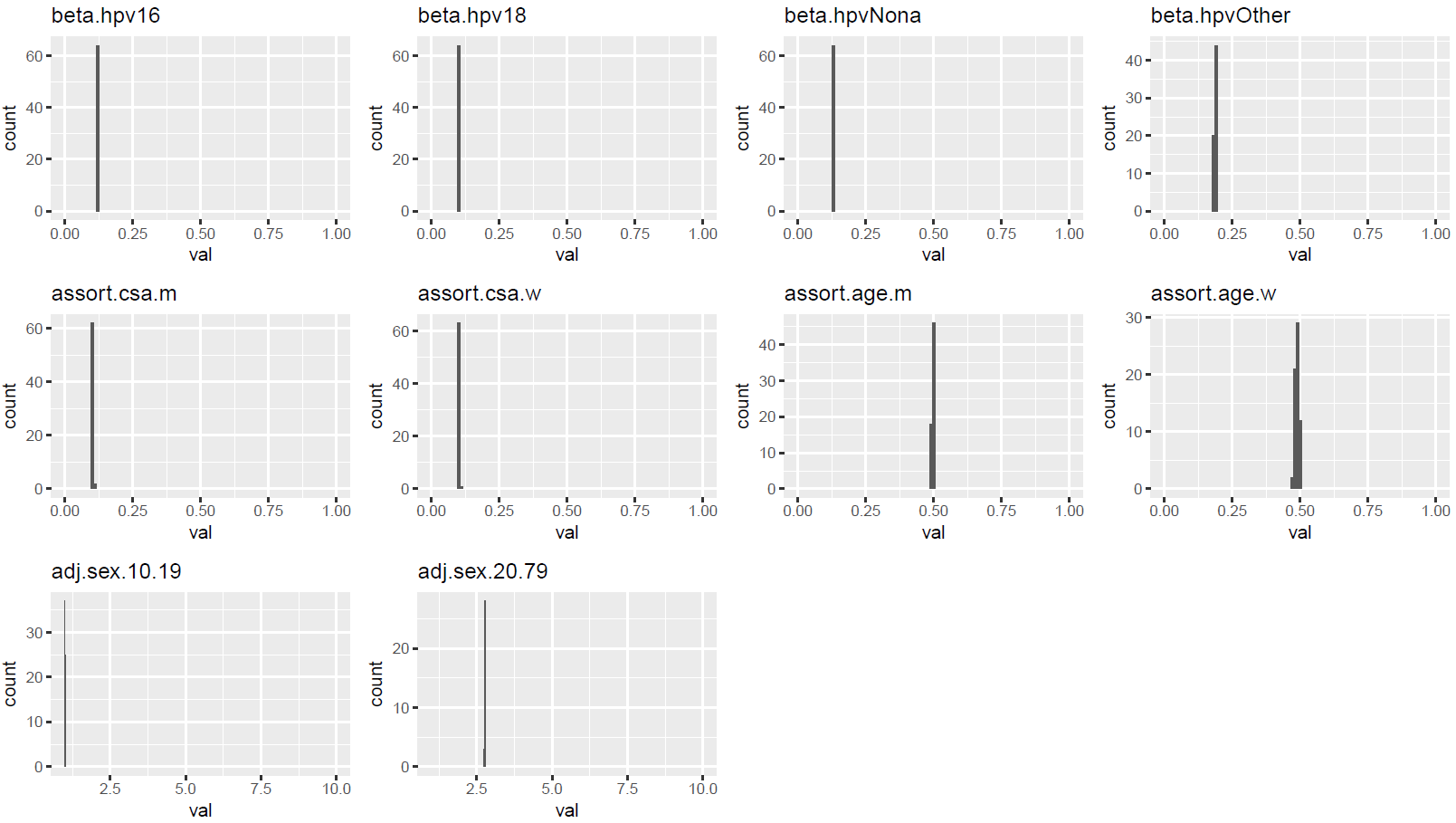
